# Sub-epidemic model forecasts for COVID-19 pandemic spread in the USA and European hotspots, February-May 2020

**DOI:** 10.1101/2020.07.03.20146159

**Authors:** Gerardo Chowell, Richard Rothenberg, Kimberlyn Roosa, Amna Tariq, James M. Hyman, Ruiyan Luo

## Abstract

Mathematical models have been widely used to understand the dynamics of the ongoing coronavirus disease 2019 (COVID-19) pandemic as well as to predict future trends and assess intervention strategies. The asynchronicity of infection patterns during this pandemic illustrates the need for models that can capture dynamics beyond a single-peak trajectory to forecast the worldwide spread and for the spread within nations and within other sub-regions at various geographic scales. Here, we demonstrate a five-parameter sub-epidemic wave modeling framework that provides a simple characterization of unfolding trajectories of COVID-19 epidemics that are progressing across the world at different spatial scales. We calibrate the model to daily reported COVID-19 incidence data to generate six sequential weekly forecasts for five European countries and five hotspot states within the United States. The sub-epidemic approach captures the rise to an initial peak followed by a wide range of post-peak behavior, ranging from a typical decline to a steady incidence level to repeated small waves for sub-epidemic outbreaks. We show that the sub-epidemic model outperforms a three-parameter Richards model, in terms of calibration and forecasting performance, and yields excellent short- and intermediate-term forecasts that are not attainable with other single-peak transmission models of similar complexity. Overall, this approach predicts that a relaxation of social distancing measures would result in continuing sub-epidemics and ongoing endemic transmission. We illustrate how this view of the epidemic could help data scientists and policymakers better understand and predict the underlying transmission dynamics of COVID-19, as early detection of potential sub-epidemics can inform model-based decisions for tighter distancing controls.

## Introduction

The asynchronicity of the infection patterns of the current coronavirus disease 2019 (COVID-19) pandemic illustrates the need for models that can capture complex dynamics beyond a single-peak trajectory to forecast the worldwide spread. This is also true for the spread within nations and within other sub-regions at various geographic scales. The infections in these asynchronous transmission networks underlie the reported infection data and need to be accounted for in forecasting models.

We analyze the COVID-19 pandemic assuming that the total number of new infections is the sum of all the infections created in multiple asynchronous outbreaks at differing spatial scales. We assume there are weak ties across sub-populations, so we represent the overall epidemic as an aggregation of *sub-epidemics*, rather than a single, universally connected outbreak. The sub-epidemics can start at different time points and affect different segments of the population in different geographic areas. Thus, we model sub-epidemics associated with transmission chains that are asynchronously triggered and that progress somewhat independently from the other sub-epidemics.

Jewell et al. (*1*) review the difficulties associated with long-term forecasting of the ongoing COVID-19 pandemic using statistical models that are not based on transmission dynamics. They also describe the limitations of models that use established mortality curves to calculate the pace of growth, the most likely inflection point, and subsequent diminution of the epidemic. The review analyzes the need for broad uncertainty bands, particularly for sub-national estimates. It also addresses the unavoidable volatility of both reporting and estimates based on reports. The analysis, delivered in the spirit of caution rather than remonstration, implies the need for other approaches that depend on overall transmission dynamics or large-scale agent-based simulations. Our sub-epidemic approach addresses this need in both the emerging and endemic stages of an epidemic.

This approach is analogous to the model used by Blower et al. (*2*) to demonstrate how the rise and endemic leveling of tuberculosis outbreaks could be explained by dynamical changes in the transmission parameters. A related multi-stage approach was used by Garnett (*3*) to explain the pattern of spread for sexually transmitted diseases and changes in the reproductive number during the course of an epidemic. Rothenberg et al. (*4*) demonstrated that the national curve of Penicillinase-Producing Neisseria gonorrhoeae occurrence resulted from multiple asynchronous outbreaks.

As with HIV/AIDS, which has now entered a phase of intractable endemic transmission in some areas (*5*), COVID-19 is likely to become endemic. New vaccines and pharmacotherapy might mitigate the transmission, but the disease will not be eradicated in the foreseeable future. Some earlier predictions based on mathematical models predicted that COVID-19 would soon disappear or approach a very low-level endemic equilibrium determined by herd immunity. To avoid unrealistic medium-range projections, some investigators artificially truncate the model projections before the model reaches these unrealistic forecasts.

Here, we demonstrate a five-parameter sub-epidemic wave modeling framework that provides a simple characterization of unfolding trajectories of COVID-19 epidemics that are progressing across the world at different spatial scales (*6*). We systematically assess calibration and forecasting performance for the ongoing COVID-19 pandemic in hotspots located in the USA and Europe using the sub-epidemic wave model, and we compare results with those obtained using the Richards model, a well-known three-parameter single-peak growth model (*7*). The sub-epidemic approach captures the rise to an initial peak followed by a wide range of post-peak behavior, ranging from a typical decline to a steady incidence level to repeated small waves for sub-epidemic outbreaks. This framework yields excellent short- and intermediate-term forecasts that are not attainable with other single-peak transmission models of similar complexity, whether mechanistic or phenomenological. We illustrate how this view of the epidemic could help data scientists and policymakers better understand and predict the underlying transmission dynamics of COVID-19.

## Methods

### Country-level data

We retrieved daily reported cumulative case data of the COVID-19 pandemic for France, the United Kingdom (UK), and the United States of America (USA) from the World Health Organization (WHO) website (*8*) and for Spain and Italy from the corresponding governmental websites (*9, 10*) from early February to May 24, 2020. We calculated the daily incidence from the cumulative trajectory and analyzed the incidence trajectory for the 5 countries.

### State-level US data

We also retrieved daily cumulative case count data from The COVID Tracking Project (*11*) from February 27, 2020 to May 24, 2020 for five representative COVID-19 hotspot states in the USA, namely New York, Louisiana, Georgia, Arizona and Washington.

### Sub-epidemic wave modeling motivation

The concept of weak ties was originally proposed by Granovetter in 1973 (*12*) to form a connection between microevents and macro events. We use this idea to link the person-to-person viral transmission of severe acute respiratory syndrome coronavirus 2 (SARS-CoV-2) to the trajectory of the COVID-19 epidemic. The transient connection between two people with different personal networks that results in the transference of the virus between the networks would be viewed as a weak tie. This event can cause asynchronous epidemic curves within the overall network. The events can spread the infection between sub-populations defined by neighborhoods, zip codes, counties, states, or countries. The resulting epidemic curve can be modeled as the sum of asynchronous sub-epidemics that reflect the movement of the virus into new populations.

In the absence of native immunity, specific viricidal treatment, or a working vaccine, our non-pharmacological preventive tools—testing, contact tracing, social separation, isolation, lockdown—are the key influences on sub-epidemic spread. The continued importation of new cases will result in low-level endemic transmission. A model based on sub-epidemic events can forecast the level of endemic spread at a steady state. This can then be used to guide intervention efforts accounting for the continued seeding of new infections.

### Sub-epidemic modeling approach

We use a five-parameter epidemic wave model that aggregates linked overlapping sub-epidemics (*6*). The strength (e.g., weak vs. strong) of the overlap determines when the next sub-epidemic is triggered and is controlled by the onset threshold parameter, *C*_*thrs*_. The incidence defines a generalized-logistic growth model (GLM) differential equation for the cumulative number of cases, *C*_*t*_, at time *t*:

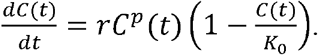

Here, *r* is the fixed growth rate, and *p is* the scaling of growth parameter, and *K*_0_ is the final size of the initial sub-epidemic. The growth rate depends on the parameter. If p=0, then the early incidence is constant over time, while if p=1 then the early incidence grows exponentially. Intermediate values of (0<p<1) describe early sub-exponential (e.g. polynomial) growth patterns.

### Model calibration and forecasting approach

For each of the ten regions, we analyzed six weekly sequential forecasts, conducted on March 30, April 6, April 13, April 20, April 27, and May 4, 2020, and assessed the calibration and forecasting performances at increasing time horizons of 2, 4, 6, …, and 20 days ahead. The models were sequentially re-calibrated each week using the most up-to-date daily curve of COVID-19 reported cases. That is, each sequential forecast included one additional week of data than the previous forecast. For comparison, we also generated forecasts using the Richards model, a well-known single-peak growth model with three parameters (*7, 13*).

## Results

### Model parameters and calibration performance

A five-parameter dynamic model, postulating sub-exponential growth in linked sub-epidemics, captures the aggregated growth curve in diverse settings (Figures 1-3 and Figures S3-S9). Using national-level data from five countries, we estimate the initial sub-exponential growth parameter (p) with a mean ranging from 0.7 to 0.9. Our analysis of five representative hotspot states in the USA indicates that early growth was sub-exponential in New York, Arizona, Georgia, and Washington (mean p ∼ 0.5-0.9) and exponential in Louisiana (Table 1). Moreover, the rate of sub-epidemic decline that captures the effects of interventions and population behavior changes is shown in Figure S10. The decay rate was fastest for Italy, followed by France, with the lowest decline rate in the USA (Table 1). Within the USA, the decline rate was the fastest for New York and Louisiana and more gradual for Georgia and Washington (Figure S10).

**Table 1.** Mean and corresponding 95% confidence intervals of estimated parameters for each COVID-19 hotspot using data up to May 24, 2020.

| **Region** | **growth rate (r)** | **Scaling of growth (p)** | **Initial sub-epidemic size (K0)** | **decline rate (q)** | **Decline function** |
| --- | --- | --- | --- | --- | --- |
| Italy | 2.5 (2.2, 2.3) | 0.69 (0.66,0.71) | 162000 (134000,182000) | 2.1 (1.61, 2.51) | Power law |
| Spain | 0.82 (0.73,0.91) | 0.87 (0.86,0.89) | 162000 (152000,172000) | 0.71 (0.64,0.79) | Exponential |
| France | 0.72 (0.43,1.4) | 0.87 (0.79, 0.92) | 101000 (83000,120000) | 1.91 (1.42,2.47) | Power law |
| UK | 0.74 (0.64,0.88) | 0.84 (0.82, 0.86) | 166000 (152000, 179000) | 0.53 (0.38,0.68) | Exponential |
| USA | 1.2 (1.0,1.3) | 0.84 (0.83,0.85) | 777000 (743000,810000) | 0.26 (0.21,0.31) | Exponential |
| New York | 3 (2.4,3.4) | 0.76 (0.74,0.79) | 173000 (160000,185000) | 0.56 (0.49,0.60) | Exponential |
| Louisiana | 0.37 (0.35,0.40) | 0.99 (0.97,1.0) | 18400 (15900,21300) | 1.20 (0.96,1.47) | Power law |
| Georgia | 0.56 (0.43,0.96) | 0.88 (0.77,0.93) | 13900 (11100,17300) | 0.20 (0.00003,0.60) | Power law |
| Arizona | 3.3 (1.9,5.1) | 0.54 (0.47, 0.63) | 16900 (13300,25000) | 0.26 (0.0004,1.2) | Exponential |
| Washington | 0.93 (0.76,1.2) | 0.79 (0.74,0.85) | 4330 (3360,5390) | 0.46 (0.21,0.64) | Power law |

**Figure 1.**
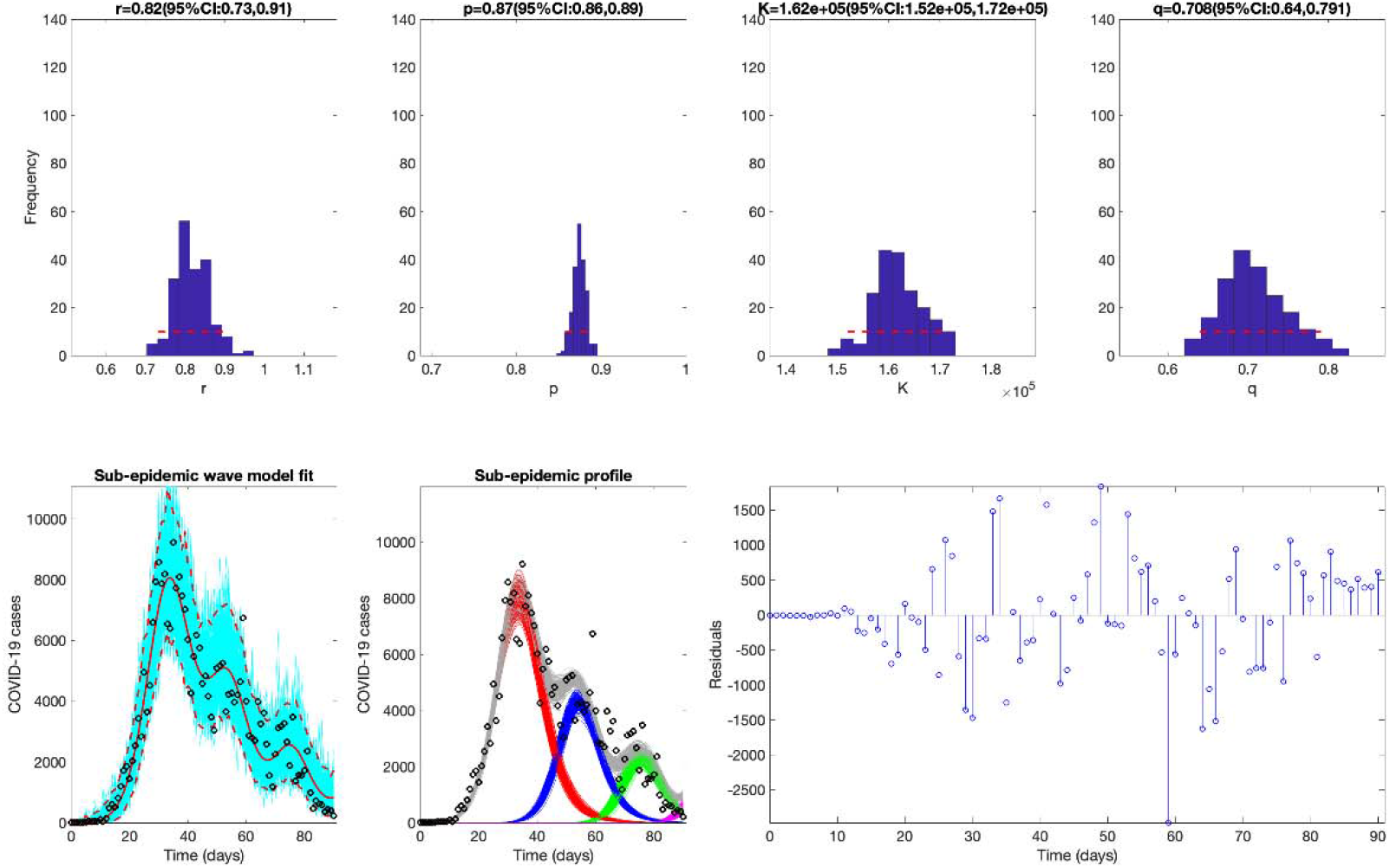
The best fit of the sub-epidemic model to the COVID-19 epidemic in Spain. The sub-epidemic wave model successfully captures the multimodal pattern of the COVID-19 epidemic. Further, parameter estimates are well identified, as indicated by their relatively narrow confidence intervals. The top panels display the empirical distribution of Bottom panels show the model fit (left), the sub-epidemic profile (center), and the residuals (right). Black circles correspond to the data points. The best model fit (solid red line) and 95% prediction interval (dashed red lines) are also shown. Cyan curves are the associated uncertainty from individual bootstrapped curves. Three hundred realizations of the sub-epidemic waves are plotted using different colors.

The calibration performance across all regions presented in Figures S1-S2 is substantially better for the overlapping sub-epidemic model compared to the Richards model based on each of the performance metrics (for MAE, MSE, and MIS, smaller is better; for 95% PI coverage, larger is better). An informative example of the model fit to the trajectory of the COVID-19 epidemic in Spain (Figure 1) shows the early growth of the epidemic in a single large sub-epidemic followed by a smaller sub-epidemic (blue in row 2, column 2 of Figure 1), which is then followed by a much smaller sub-epidemic (green). In row 1 (Figure 1), the parameter distributions demonstrate relatively small confidence intervals. Thus, the model captures a common phenomenon in epidemic situations: an initial steep rise, followed by a leveling or decline, then a second rise, and a subsequent repeat of the same pattern. A somewhat different pattern is observed in the USA, which experienced sustained transmission with high mortality for a long period (Figure 2). A single epidemic wave failed to capture the early growth phase and the later leveling off; whereas, the aggregation of multiple sub-epidemics produces a better fit to the observed dynamics. In comparison, New York, the early epicenter of the pandemic in the USA, displays a similar sub-epidemic profile, while the sub-epidemic sizes decline at a much faster rate (Figure 3).

**Figure 2.**
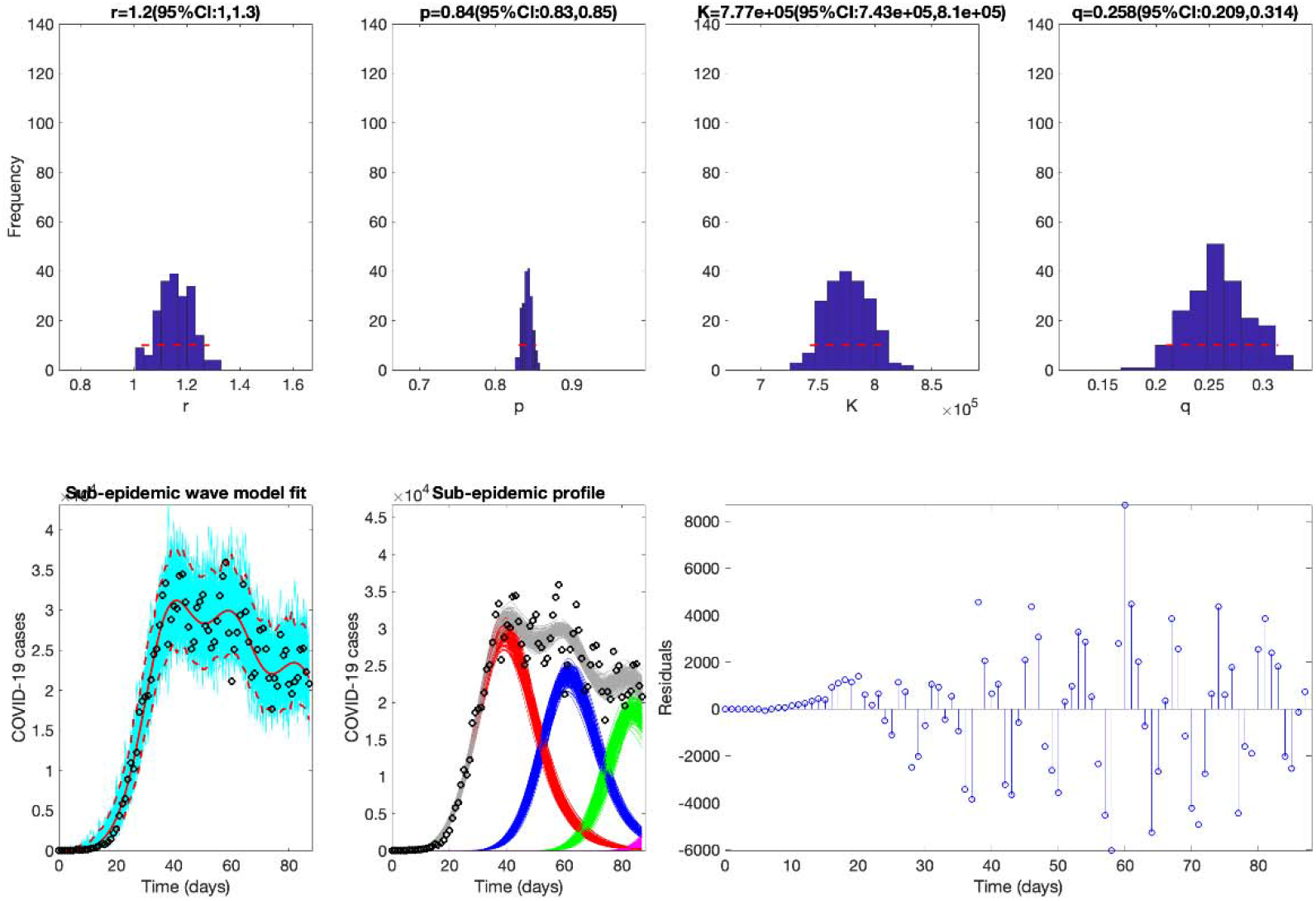
The best fit of the sub-epidemic model to the COVID-19 epidemic in the USA. The sub-epidemic wave model successfully captures the multimodal pattern of the COVID-19 epidemic. Further, parameter estimates are well identified, as indicated by their relatively narrow confidence intervals. The top panels display the empirical distribution of Bottom panels show the model fit (left), the sub-epidemic profile (center), and the residuals (right). Black circles correspond to the data points. The best model fit (solid red line) and 95% prediction interval (dashed red lines) are also shown. Cyan curves are the associated uncertainty from individual bootstrapped curves. Three hundred realizations of the sub-epidemic waves are plotted using different colors.

**Figure 3.**
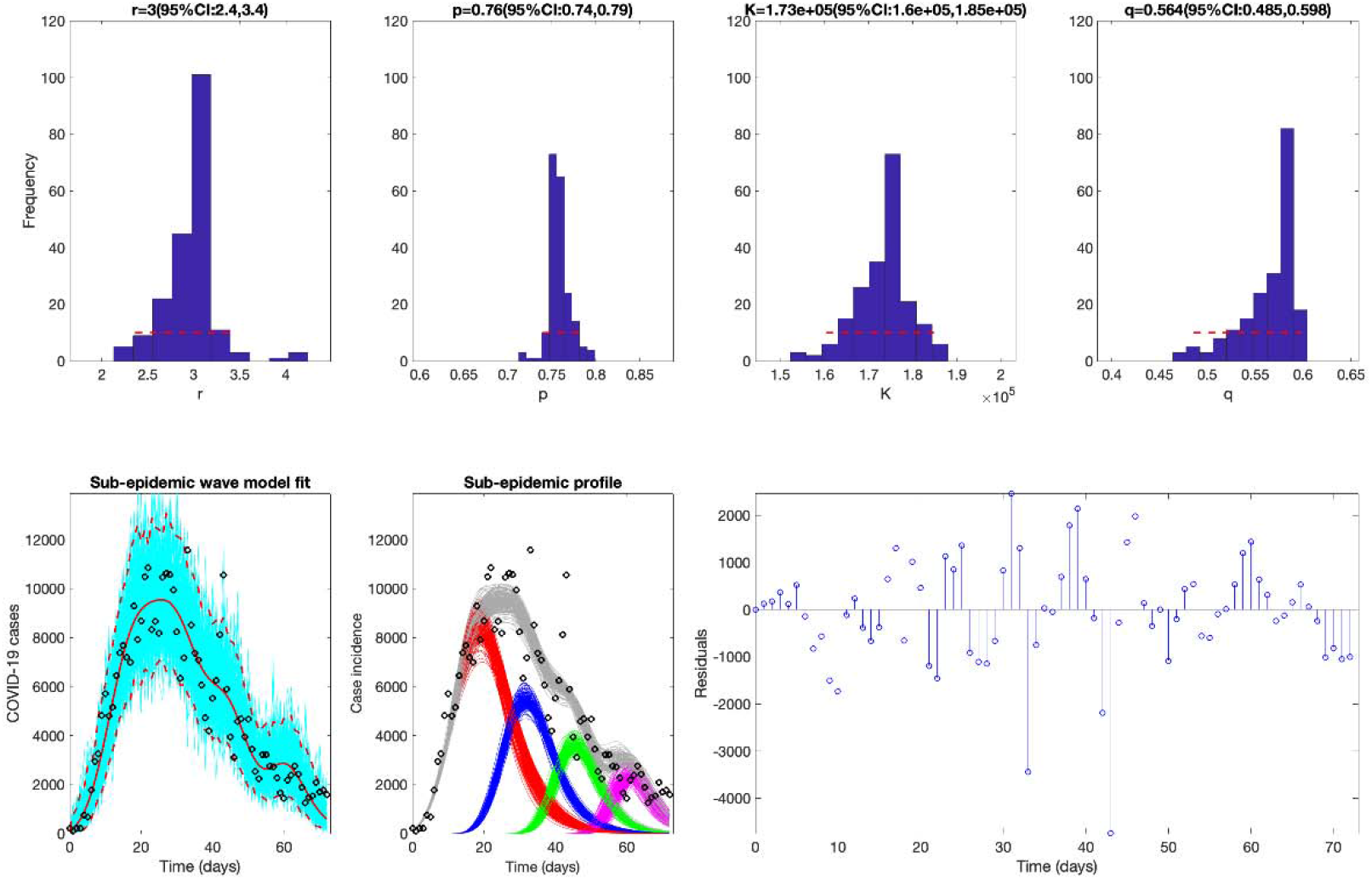
The best fit of the sub-epidemic model to the COVID-19 epidemic in New York State. The sub-epidemic wave model successfully captures the overlapping sub-epidemic growth pattern of the COVID-19 epidemic. Further, parameter estimates are well identified, as indicated by their relatively narrow confidence intervals. The top panels display the empirical distribution of Bottom panels show the model fit (left), the sub-epidemic profile (center), and the residuals (right). Black circles correspond to the data points. The best model fit (solid red line) and 95% prediction interval (dashed red lines) are also shown. Cyan curves are the associated uncertainty from individual bootstrapped curves. Three hundred realizations of the sub-epidemic waves are plotted using different colors.

Similar composite figures for the remaining regions (Figures S3-S9) demonstrate diverse patterns of underlying sub-epidemic waves. For example, Italy experienced a single peak, largely the result of an initial sub-epidemic (in red), that was quickly followed by several rapidly declining sub-epidemics that slowed the downward progression (Figure S3). The UK’s sub-epidemic profile resembles that of the USA, but the sub-epidemics decline at a faster rate (Figure S5; Table 1).

### Forecasting performance

The sub-epidemic wave model outperformed the simpler Richards model in most of the 2-20 day ahead forecasts (see Figure 4 and Figures S11-S19). We observe that the sub-epidemic model forecasting accuracy increases as evidence for the second sub-epidemic appears in the data. For instance, the initial forecasts for the USA using the sub-epidemic model (Figures 5 & S20) underestimate reported incidence for the 20 days after April 7th, which is likely attributable to the unexpected leveling off of the epidemic wave. However, this model provided more accurate forecasts in subsequent 20-day forecasts.

**Figure 4.**
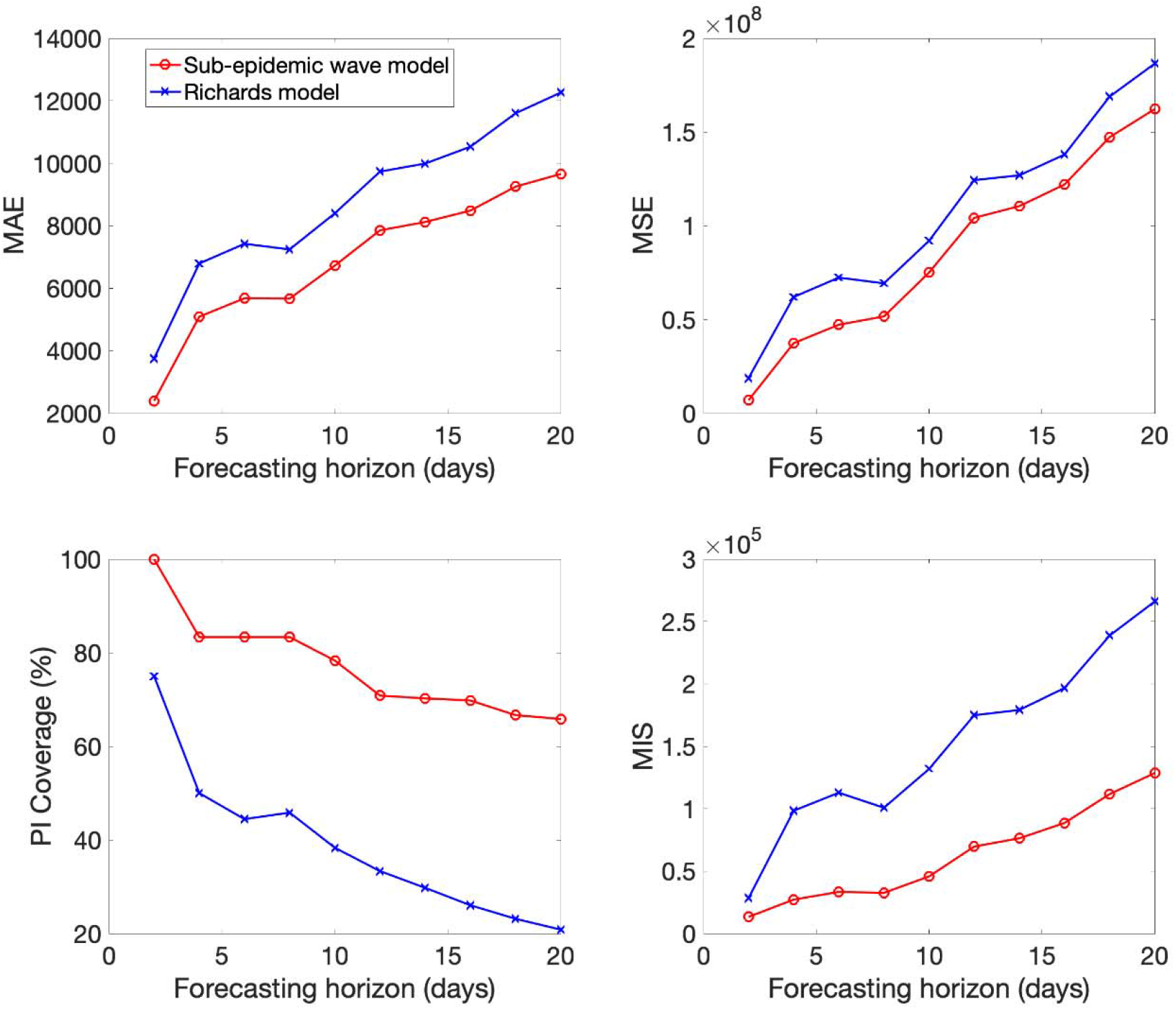
Mean performance of the sub-epidemic wave (red) and the Richards (blue) models in 2-20 day ahead forecasts conducted during the epidemic in the USA. The sub-epidemic model outperformed the Richards model across all metrics and forecasting horizons.

**Figure 5.**
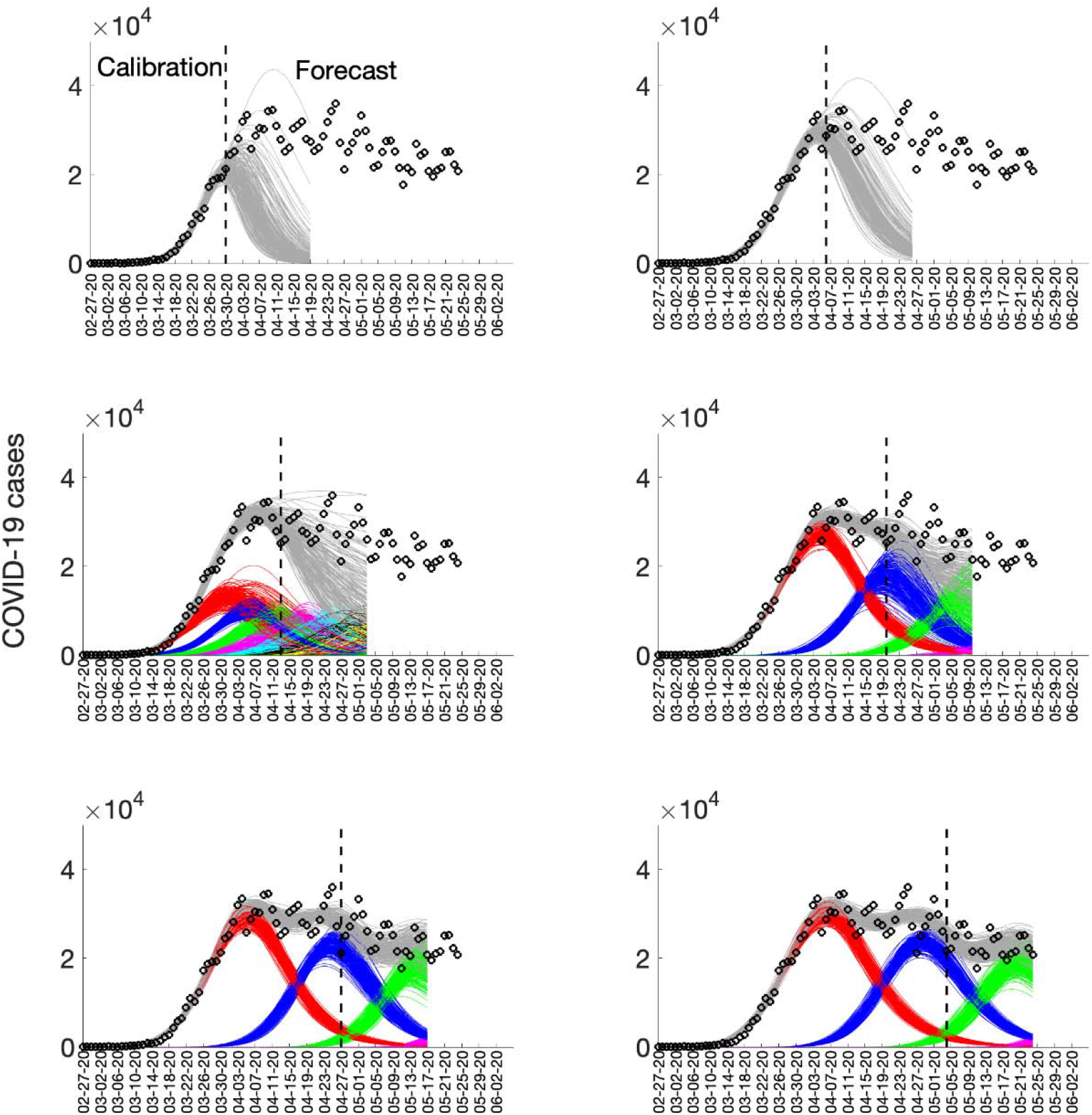
Sub-epidemic profiles of the sequential 20-day ahead forecasts for the COVID-19 epidemic in the USA. Different colors represent different sub-epidemics of the epidemic wave profile. The aggregated trajectories are shown in gray, and black circles correspond to the data points. The vertical line separates the calibration period (left) from the forecasting period (right). The sequential forecasts were conducted on March 30, April 6, April 13, April 20, April 27, and May 4, 2020.

Similarly, sub-national models of the USA state trajectories confirm the general findings of fit and 20-day forecasting (see supplementary materials). Among the most striking of these is the sub-epidemic structure modeled for New York state (Figure S25). When the sub-epidemic model is calibrated by April 7, 2020, a single sub-epidemic is observed; however, subsequent weeks of data helped infer an underlying overlapping sub-epidemic structure and correctly forecasted the subsequent downward trend. With variation, other states shown in the supplementary materials provided similar confirmation of the method.

## Discussion

Our sub-epidemic modeling framework is based on the premise that the aggregation of regular sub-epidemic dynamics can determine the shape of the trajectory of epidemic waves observed at larger spatial scales. This framework has been particularly suitable for forecasting the spatial wave dynamics of the COVID-19 pandemic, where the trajectory of the epidemic at different spatial scales does not display a single peak followed by a “burnout” period, but instead follows more complex transmission patterns including leveling off, plateaus, and long-tail decline periods. The model overwhelmingly outperformed a standard growth model that only allows for single-peak transmission dynamics. Model parameters also inform the effect of interventions and population behavior changes in terms of the sub-epidemic decay rate.

Overall, this approach predicts that a relaxation of the tools currently at our disposal— primarily aimed at preventing person-to-person and person-to-surface contact—would result in continuing sub-epidemics and ongoing endemic transmission. If we add widespread availability of testing, contact tracing, and cluster investigation (e.g. nursing homes, meatpacking plants, and other sites of unavoidable congregation), early suppression of sub-epidemics may be possible. The United States leads in the total number of tests performed, but it is currently ranked 25th among all nations in testing per capita (*14*). The sub-epidemic description of COVID-19 transmission provides a rationale for substantial increases in testing.

Parsimony in model construction is not an absolute requirement, but it has several advantages. With fewer parameters to estimate, the joint simulations are more efficient and more understandable. Degenerate results are more easily avoided, and, when properly constructed, confidence intervals for the key parameters are more constrained. In our projections, we fit five parameters to the data:

1. The onset threshold parameter, *C*_*thr*_, that triggers the onset of a new sub-epidemic and determines if the overlap is weak or strong,
2. The new epidemic starting size, *K*_0_,
3. The size of consecutive sub-epidemics decline rates *q*,
4. The positive parameter *r* denoting the growth rate of a sub-epidemic, and
5. The “scaling of growth” parameter *p* ∈[0,1] (exponential or sub-exponential).

As shown in Figure 1, the confidence limits for these parameters are narrow, and the scaling of growth parameter is constantly in the 0.8 to 0.9 range (Table 1).

Short-term forecasting is an important attribute of the model. Though long-term forecasts are of value, their dependability varies inversely with the time horizon. The 20-day forecasts are most valuable for the monitoring, management, and relaxation of the social distancing requirements. The early detection of potential sub-epidemics can signal the need for strict distancing controls, and the reports of cases can identify the geographic location of incubating sub-epidemics. No single model or method can provide an unerring approach to epidemic control. The multiplicity of models now available can be viewed as a source of confusion, but it is better thought of as a strength that provides multiple perspectives. The sub-epidemic approach adds to the current armamentarium for guiding us through the COVID-19 pandemic.

## Data Availability

All data are publicly available

https://www.who.int/emergencies/diseases/novel-coronavirus-2019/situation-reports

https://cnecovid.isciii.es/

https://github.com/pcm-dpc/COVID-19

https://covidtracking.com/data

## Author contributions

GC conceived the study. KR and AT contributed to data analysis. All authors contributed to the interpretation of the results. GC and RR wrote the first draft of the manuscript. All authors contributed to writing subsequent drafts of the manuscript. All authors read and approved the final manuscript.

## Data and materials availability

All data are publicly available.

## Supplementary Materials for

### Sub-epidemic modeling approach

We use a five-parameter epidemic wave model that aggregates linked overlapping sub-epidemics (*1*). The strength (e.g., weak vs. strong) of the overlap determines when the next sub-epidemic is triggered and is controlled by the onset threshold parameter,*C*_*thrs*_. The incidence defines a generalized-logistic growth model (GLM) differential equation for the cumulative number of cases,*C*_*t*_, at time *t*:

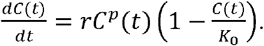

The sub-epidemics are modeled by a system of coupled differential equations:

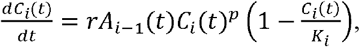

Here *C*_*i*_(*t*) is the cumulative number of infections for sub-epidemic *i* and *K*_*i*_ is the size of the *i*_*th*_ sub-epidemic where *i* = 1, …, *n*. Starting from an initial sub-epidemic size *K*_0_, the size of consecutive sub-epidemics *K*_*i*_ decline at the rate *q* following an exponential or power-law function.

The onset timing of the (*i* + 1)_*th*_ sub-epidemic is determined by the indicator variable *A*_*i*_ (*t*). This results in a coupled system of sub-epidemics where the (*i* + 1)_*th*_ sub-epidemic is triggered when the cumulative number of cases for the *i*_*th*_ sub-epidemic exceeds a total of *C*_*thr*_ cases. The sub-epidemics are *overlapping* because the (*i* + 1)_*th*_ sub-epidemic takes off before the *i*_*th*_ sub-epidemic completes its course. That is,

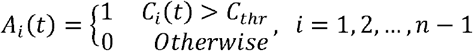

The threshold parameters are defined so 1 ≤ *C*_*thr*_ < *K*_0_ and *A*_0_ (*t*) for the first sub-epdemic.

This framework allows the size of the *i*_*th*_ sub-epidemic (*K*_*i*_) to remain steady or decline based on the factors underlying the transmission dynamics. These factors could include a gradually increasing effect of public health interventions or population behavior changes that mitigate transmission. We consider both exponential and inverse decline functions to model the size of consecutive sub-epidemics.

### Exponential decline of sub-epidemic sizes

If consecutive sub-epidemics decline exponentially, then *K*_*i*_ is given by:

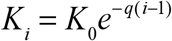

Where *K*_0_ is the size of the initial sub-epidemic (*K*_1_ = *K*_0_). If *q* = 0, then the model predicts an epidemic wave comprising sub-epidemics of the same size. When *q* > 0, then the total number of sub-epidemics *n*_*tot*_ is finite and depends on *C*_*thr*_, *q*, and, *K*_0_. The sub-epidemic is only triggered if *C*_*thr*_ ≤ *K*_*i*_, resulting in a finite number of sub-epidemics,

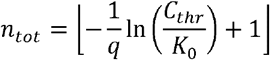

The brackets ⌊ * ⌋ denote the largest integer that is smaller than or equal to *. The total size of the epidemic wave composed of *n*_*tot*_ overlapping sub-epidemics has a closed-form solution:

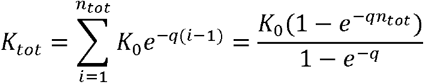

### Inverse decline of sub-epidemic sizes

The consecutive sub-epidemics decline according to the inverse function given by:

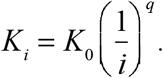

When *q* > 0, then the total number of sub-epidemics *n*_*tot*_ is finite and is given by:

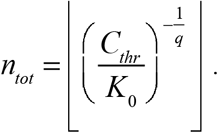

The total size of an epidemic wave is the sum of *n* overlapping sub-epidemics,

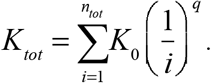

In the absence of control interventions or behavior change (*q* = 0), the total epidemic size depends on a given number *n* of sub-epidemics,

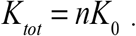

The initial number of cases is given by *C*_1_(0) = *I*_0_ where *I*_0_ is the initial number of cases in the observed case data. The cumulative cases, *C*(*t*), is the sum of all cumulative infections over the *n* overlapping sub-epidemics waves:

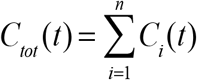

### Parameter estimation

Fitting the model to the time series of case incidence requires estimating up to five model parameters Θ= (*C*_*thr*_, *q,r, p, K*). If a single sub-epidemic is sufficient to fit the data, then the model is simplified to the three-parameter generalized-logistic growth model. The model parameters were estimated by a nonlinear least square fit of the model solution to the observed incidence data (*2*). This is achieved by searching for the set of parameters 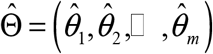 that minimizes the sum of squared differences between the observed incidence Data 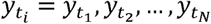 and the corresponding mean incidence curve denoted by *f* (*t*_*i*_, Θ). That is, the parameters are estimated by

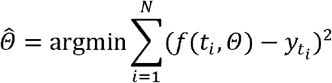

where *t*_*i*_ are the time points at which the time-series data are observed, and *N* is the number of data points available for inference. Hence, the model solution 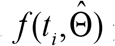 yields the best fit to the time series data 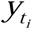, where 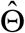 is the vector of parameter estimates.

We solve the nonlinear least squares problem using the trust-region reflective algorithm. We used parametric bootstrap, assuming an error structure described in the next section, to quantify the uncertainty in the parameters obtained by a non-linear least squares fit of the data, as described in refs. (*3, 4*). Our best-fit model solution is given by 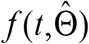 where 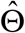 is the vector of parameter estimates. Our MATLAB (The MathWorks, Inc) code for model fitting along with outbreak datasets is publicly available (*5*).

The confidence interval for each estimated parameter and 95% prediction intervals of the model fits were obtained using parametric bootstrap (*4*). Let *S* denote the number of bootstrap realizations and 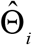 denote the re-estimation of parameter set Θ from the i^th^ bootstrap sample. The variance and confidence interval for 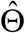 are estimated from 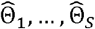. Similarly, the uncertainty of the model forecasts, 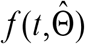, is estimated using the variance of the parametric bootstrap samples

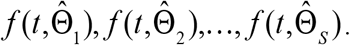

where 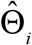 denotes the estimation of parameter set Θ from the i_th_ bootstrap sample. The 95% prediction intervals of the forecasts in the examples are calculated from the 2.5% and 97.5% percentiles of the bootstrap forecasts.

### Error structure

We model a negative binomial distribution for the error structure and assume a constant variance/mean ratio over time (i.e., the overdispersion parameter). To estimate this constant ratio, we group every four daily observations into a bin across time, calculate the mean and variance for each bin, and then estimate a constant variance/mean ratio by calculating the average of the variance/mean ratios over these bins. Exploratory analyses indicate that this ratio is frequently stable across bins, except for 1-2 extremely large values, which could result from a sudden increase or decrease in the number of reported cases. These sudden changes could result from changes in case definition or a weekend effect whereby the number of reported cases decreases systematically during weekends. Hence, these extreme large values of variance/mean ratio are excluded when estimating the constant variance/mean ratio.

### Model performance

To assess both the quality of the model fit and the short-term forecasts, we used four performance metrics: the mean absolute error (MAE), the mean squared error (MSE), the coverage of the 95% prediction intervals, and the mean interval score (MIS) (*6*). The *mean absolute error* (MAE) is given by:

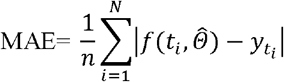

Here 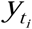 is the time series of incident cases describing the epidemic wave where *t*_*i*_ are the time points of the time series data (*7*). Similarly, the *mean squared error* (MSE) is given by:

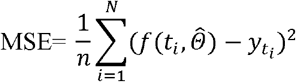

In addition, we assessed the *coverage of the 95% prediction interval*, e.g., the proportion of the observations that fell within the 95% prediction interval as well as a metric that addresses the width of the 95% prediction interval as well as coverage via the *mean interval score* (MIS) (*6, 8*) which is given by:

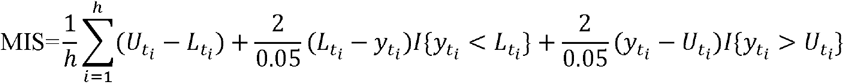

where *L*_*t*_ and *U*_*t*_ are the lower and upper bounds of the 95% prediction interval and **I**{} is an indicator function. Thus, this metric rewards for narrow 95% prediction intervals and penalizes at the points where the observations are outside the bounds specified by the 95% prediction interval where the width of the prediction interval adds up to the penalty (if any) (*6*).

The mean interval score (MIS) and the coverage of the 95% prediction intervals take into account the uncertainty of the predictions whereas the mean absolute error (MAE) and mean squared error (MSE) only assess the closeness of the mean trajectory of the epidemic to the observations (*9*). These performance metrics have also been adopted in international forecasting competitions (*8*).

For comparison purposes, we compare the performance of the sub-epidemic wave model with that obtained from the 3-parameter Richards model (*10*), a well-known single-peak growth model given by:

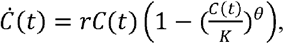

where θ determines the deviation from symmetry, and again *r* is the growth rate, and *K* is the final epidemic size.

---

**Figure S1.**
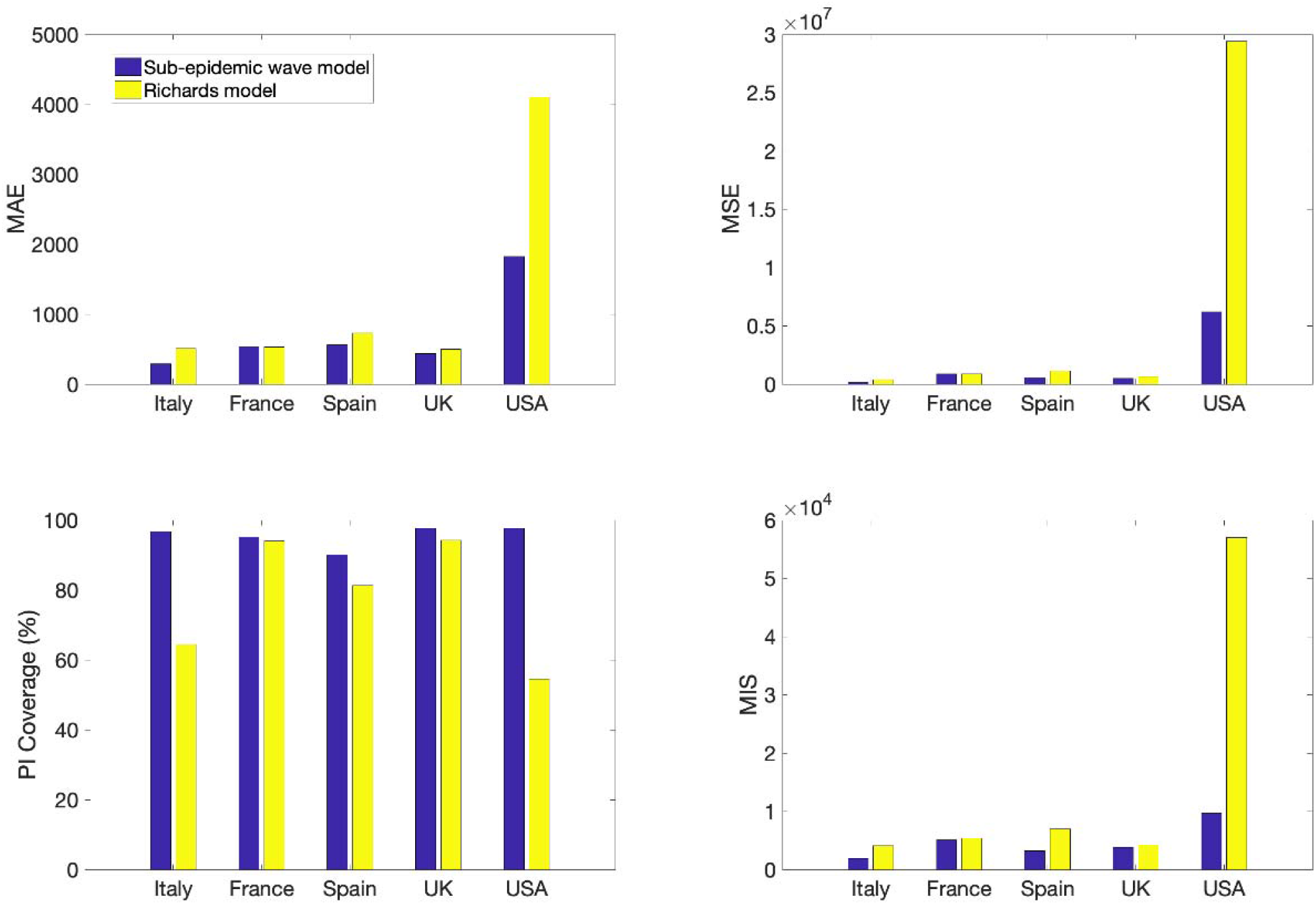
The calibration performance metrics across five countries are uniformly better for the overlapping sub-epidemic models (for MAE, MSE, and MIS, smaller is better; for % covered, larger is better).

**Figure S2.**
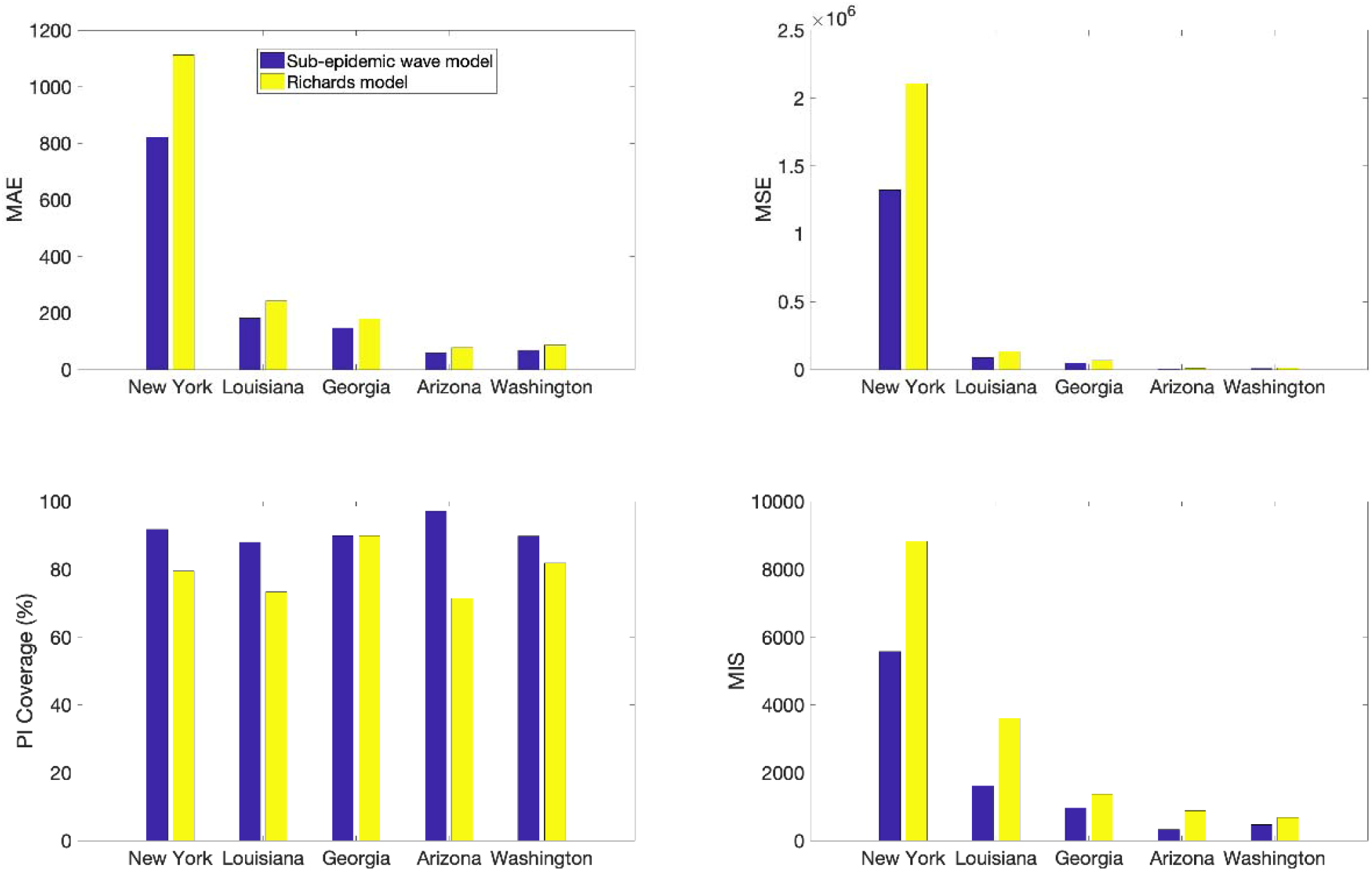
The calibration performance metrics across five hotspots in the USA are uniformly better for the overlapping sub-epidemic models (for MAE, MSE, and MIS, smaller is better; for % covered, larger is better).

**Figure S3.**
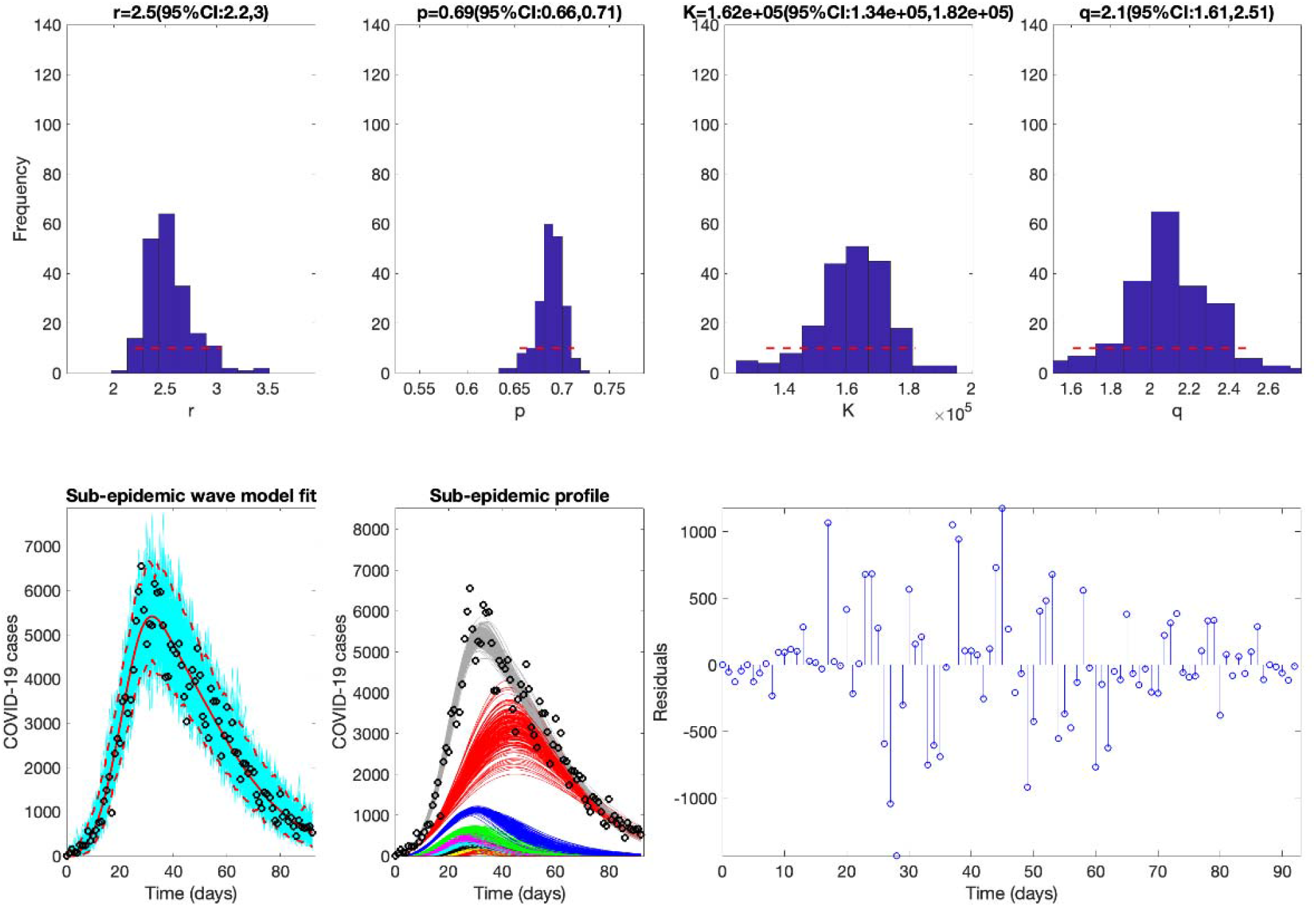
The best fit of the sub-epidemic model to the COVID-19 epidemic in Italy. The sub-epidemic wave model successfully captures the multimodal pattern of the COVID-19 epidemic. Further, parameter estimates are well identified, as indicated by their relatively narrow confidence intervals. The top panels display the empirical distribution of *r, p, K*, and *q*. Bottom panels show the model fit (left), the sub-epidemic profile (center), and the residuals (right). Black circles correspond to the data points. The best model fit (solid red line) and 95% prediction interval (dashed red lines) are also shown. Cyan curves are the associated uncertainty from individual bootstrapped curves. Three hundred realizations of the sub-epidemic waves are plotted using different colors.

**Figure S4.**
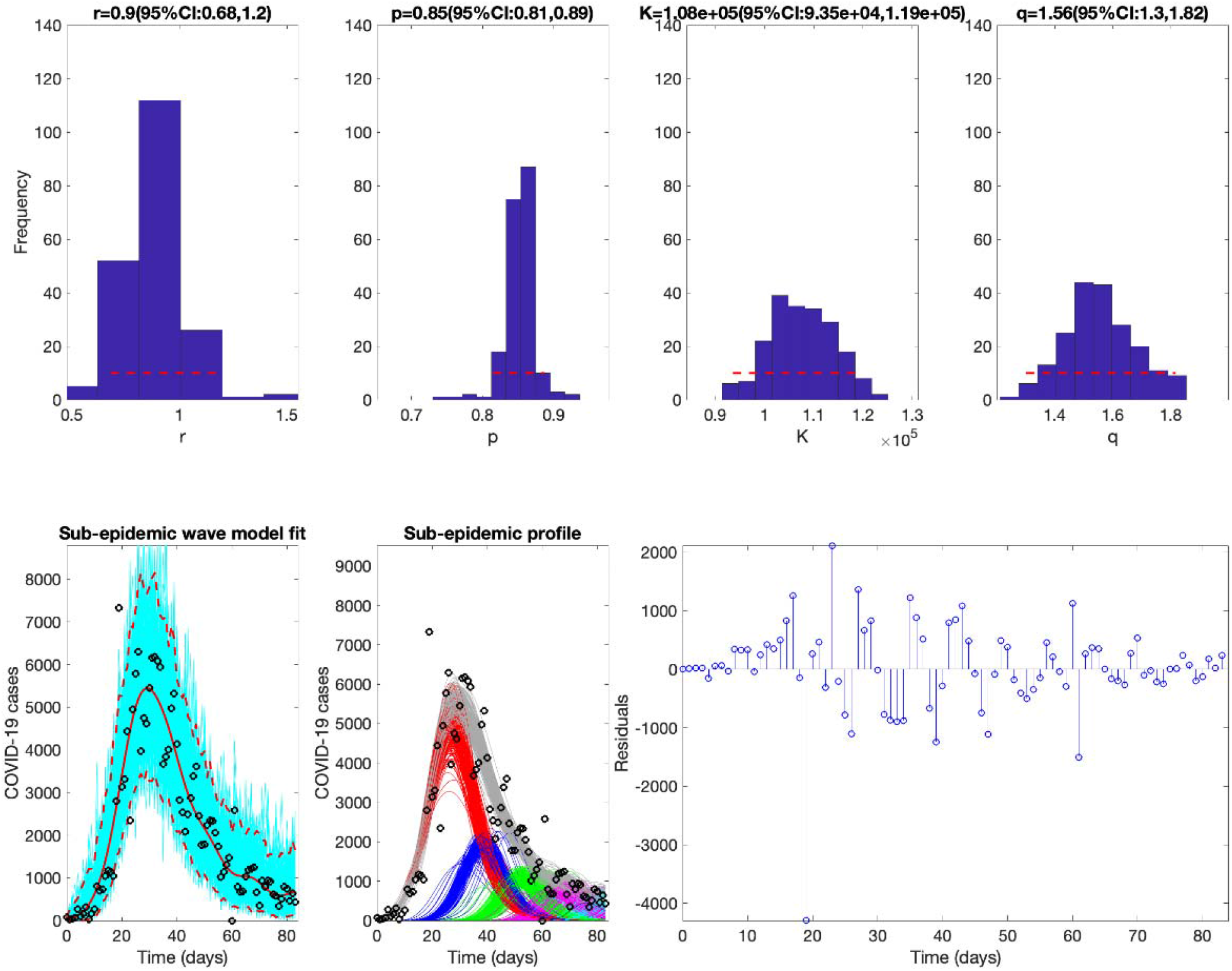
The best fit of the sub-epidemic model to the COVID-19 epidemic in France. The sub-epidemic wave model successfully captures the multimodal pattern of the COVID-19 epidemic. Further, parameter estimates are well identified, as indicated by their relatively narrow confidence intervals. The top panels display the empirical distribution of *r, p, K*, and *q*. Bottom panels show the model fit (left), the sub-epidemic profile (center), and the residuals (right). Black circles correspond to the data points. The best model fit (solid red line) and 95% prediction interval (dashed red lines) are also shown. Cyan curves are the associated uncertainty from individual bootstrapped curves. Three hundred realizations of the sub-epidemic waves are plotted using different colors.

**Figure S5.**
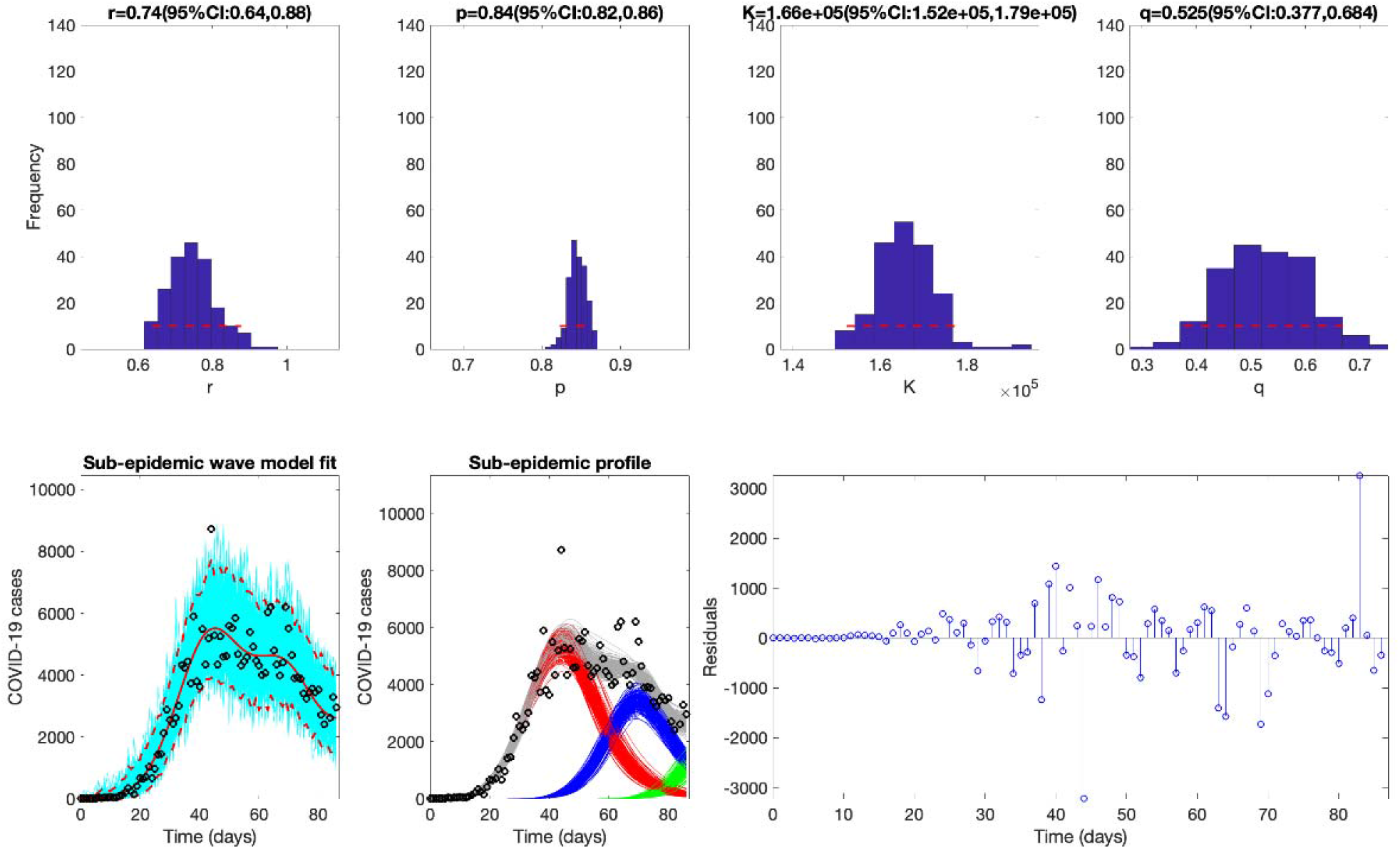
The best fit of the sub-epidemic model to the COVID-19 epidemic in the United Kingdom. The sub-epidemic wave model successfully captures the multimodal pattern of the COVID-19 epidemic. Further, parameter estimates are well identified, as indicated by their relatively narrow confidence intervals. The top panels display the empirical distribution of *r, p, K*, and *q*. Bottom panels show the model fit (left), the sub-epidemic profile (center), and the residuals (right). Black circles correspond to the data points. The best model fit (solid red line) and 95% prediction interval (dashed red lines) are also shown. Cyan curves are the associated uncertainty from individual bootstrapped curves. Three hundred realizations of the sub-epidemic waves are plotted using different colors.

**Figure S6.**
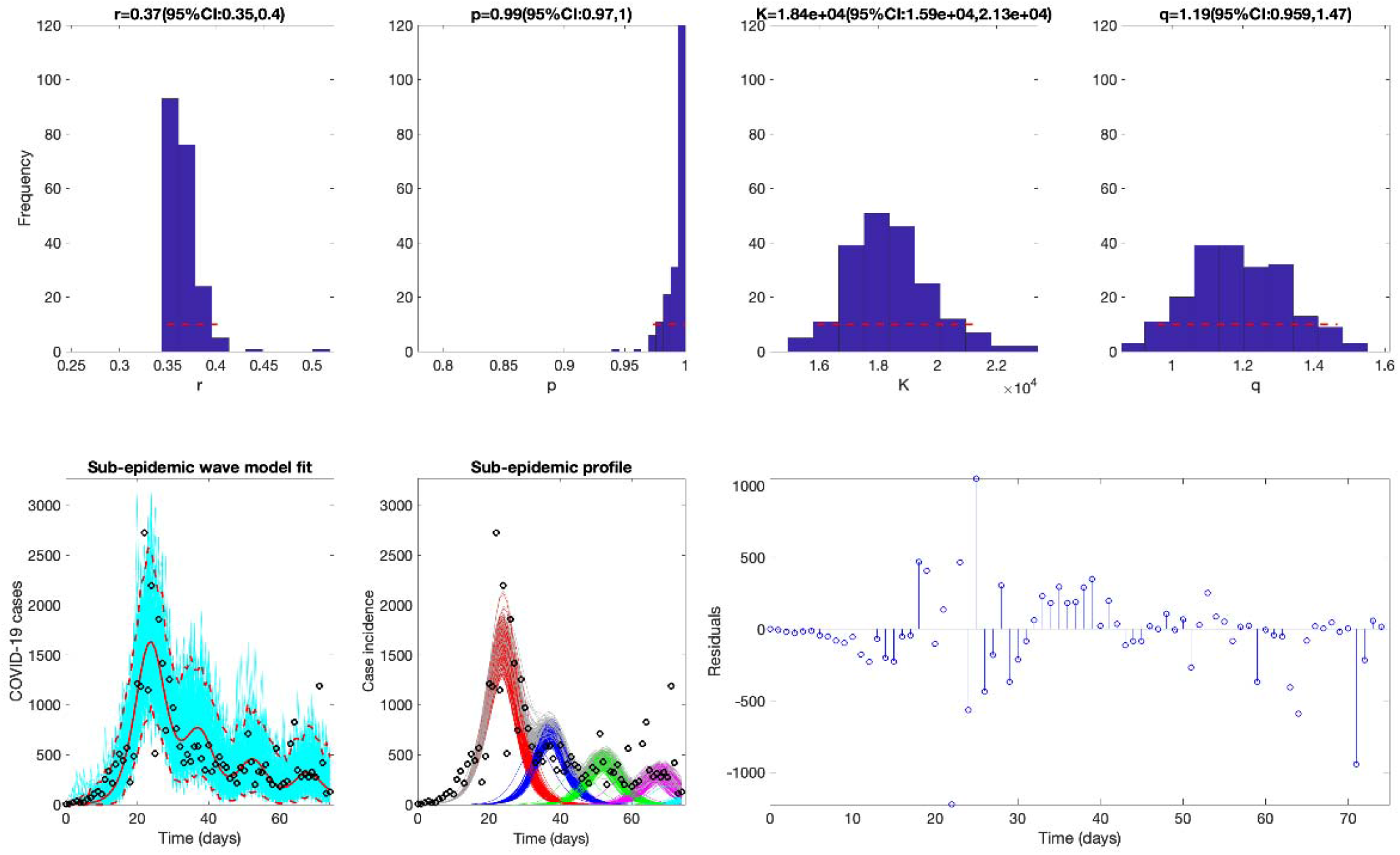
The best fit of the sub-epidemic model to the COVID-19 epidemic in Louisiana, USA. The sub-epidemic wave model successfully captures the multimodal pattern of the COVID-19 epidemic. Further, parameter estimates are well identified, as indicated by their relatively narrow confidence intervals. The top panels display the empirical distribution of *r, p, K*, and *q*. Bottom panels show the model fit (left), the sub-epidemic profile (center), and the residuals (right). Black circles correspond to the data points. The best model fit (solid red line) and 95% prediction interval (dashed red lines) are also shown. Cyan curves are the associated uncertainty from individual bootstrapped curves. Three hundred realizations of the sub-epidemic waves are plotted using different colors.

**Figure S7.**
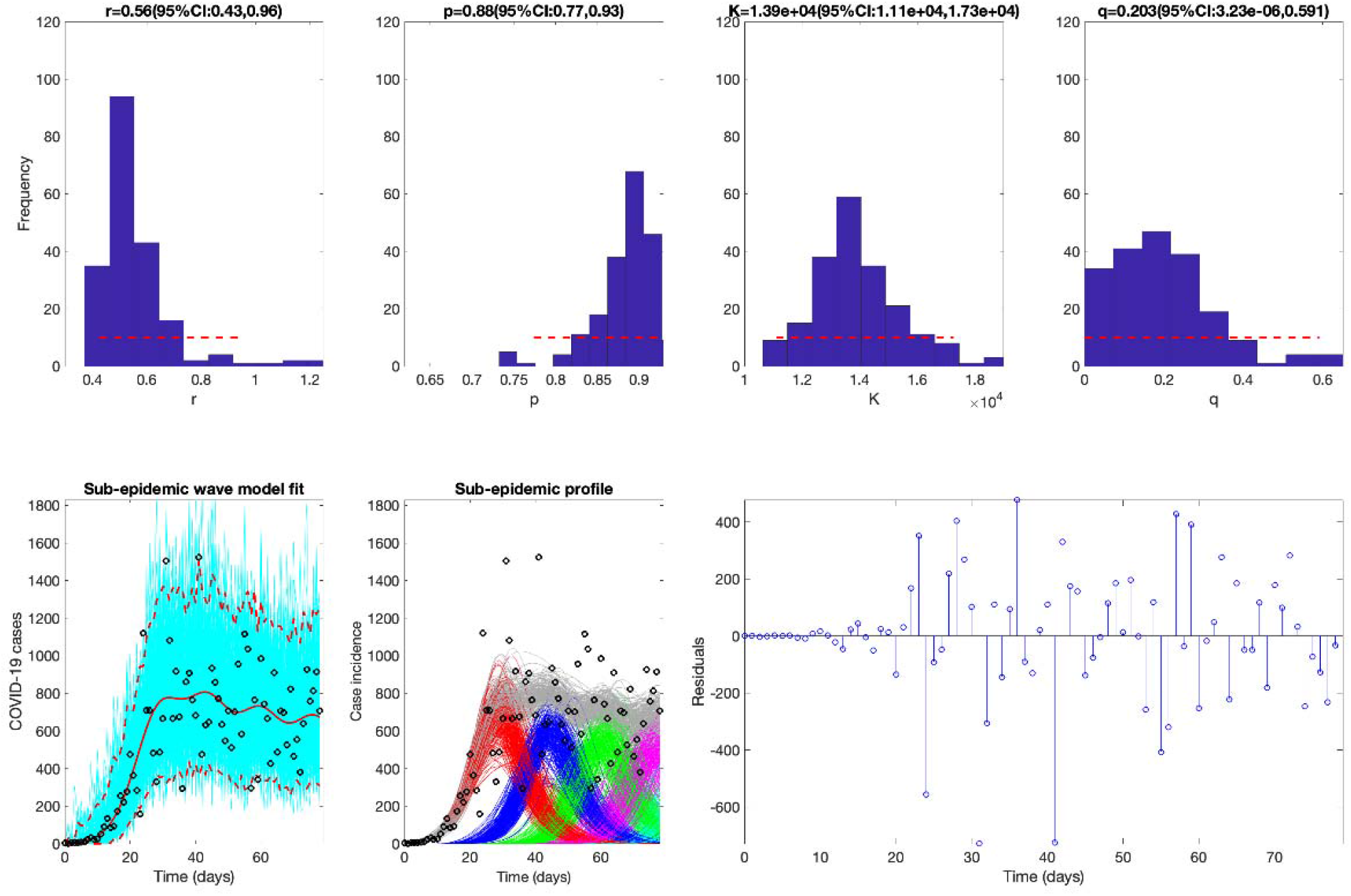
The best fit of the sub-epidemic model to the COVID-19 epidemic in Georgia, USA. The sub-epidemic wave model successfully captures the multimodal pattern of the COVID-19 epidemic. Further, parameter estimates are well identified, as indicated by their relatively narrow confidence intervals. The top panels display the empirical distribution of *r, p, K*, and *q*. Bottom panels show the model fit (left), the sub-epidemic profile (center), and the residuals (right). Black circles correspond to the data points. The best model fit (solid red line) and 95% prediction interval (dashed red lines) are also shown. Cyan curves are the associated uncertainty from individual bootstrapped curves. Three hundred realizations of the sub-epidemic waves are plotted using different colors.

**Figure S8.**
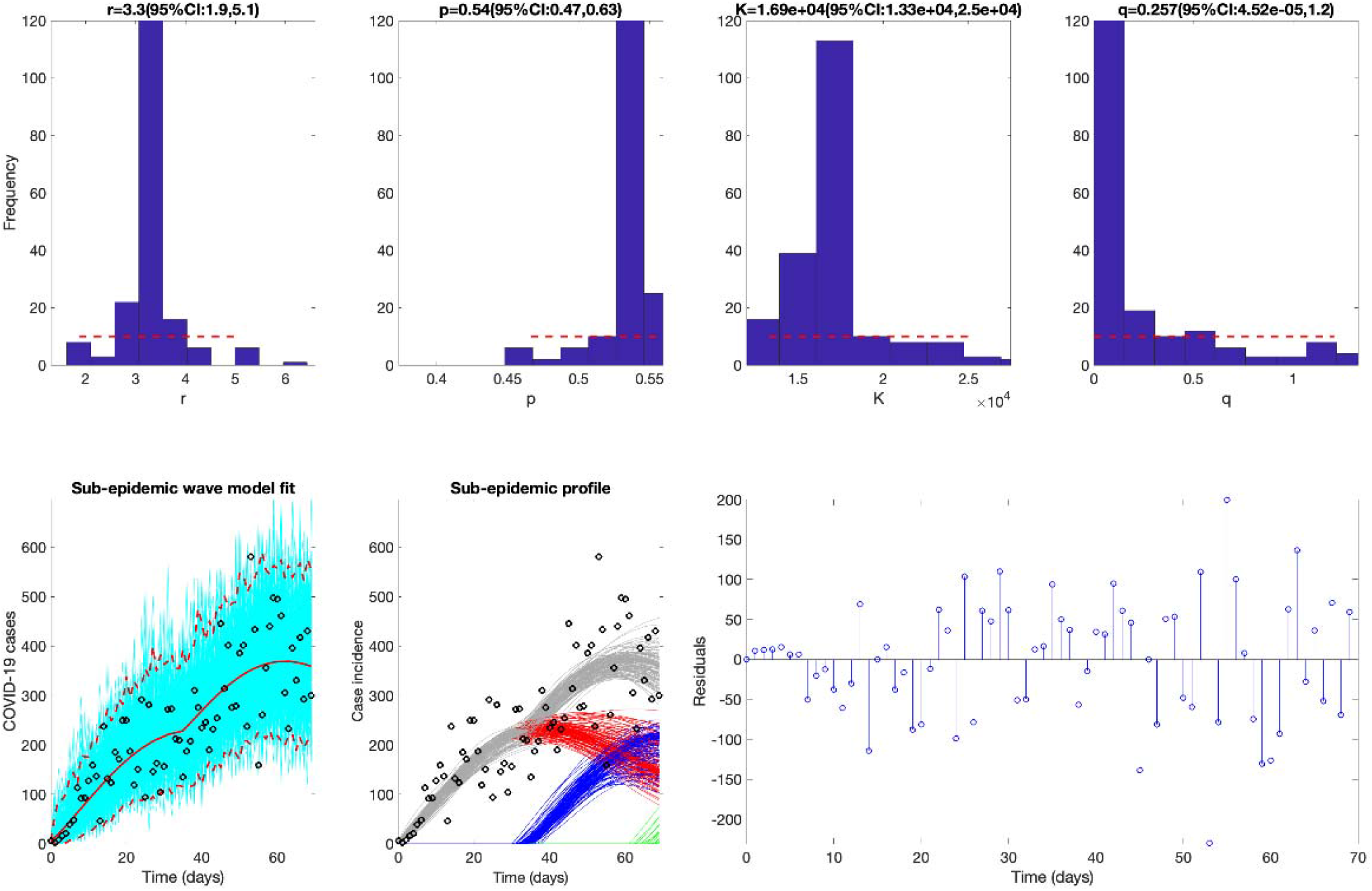
The best fit of the sub-epidemic model to the COVID-19 epidemic in Arizona, USA. The sub-epidemic wave model successfully captures the multimodal pattern of the COVID-19 epidemic. Further, parameter estimates are well identified, as indicated by their relatively narrow confidence intervals. The top panels display the empirical distribution of *r, p, K*, and *q*. Bottom panels show the model fit (left), the sub-epidemic profile (center), and the residuals (right). Black circles correspond to the data points. The best model fit (solid red line) and 95% prediction interval (dashed red lines) are also shown. Cyan curves are the associated uncertainty from individual bootstrapped curves. Three hundred realizations of the sub-epidemic waves are plotted using different colors.

**Figure S9.**
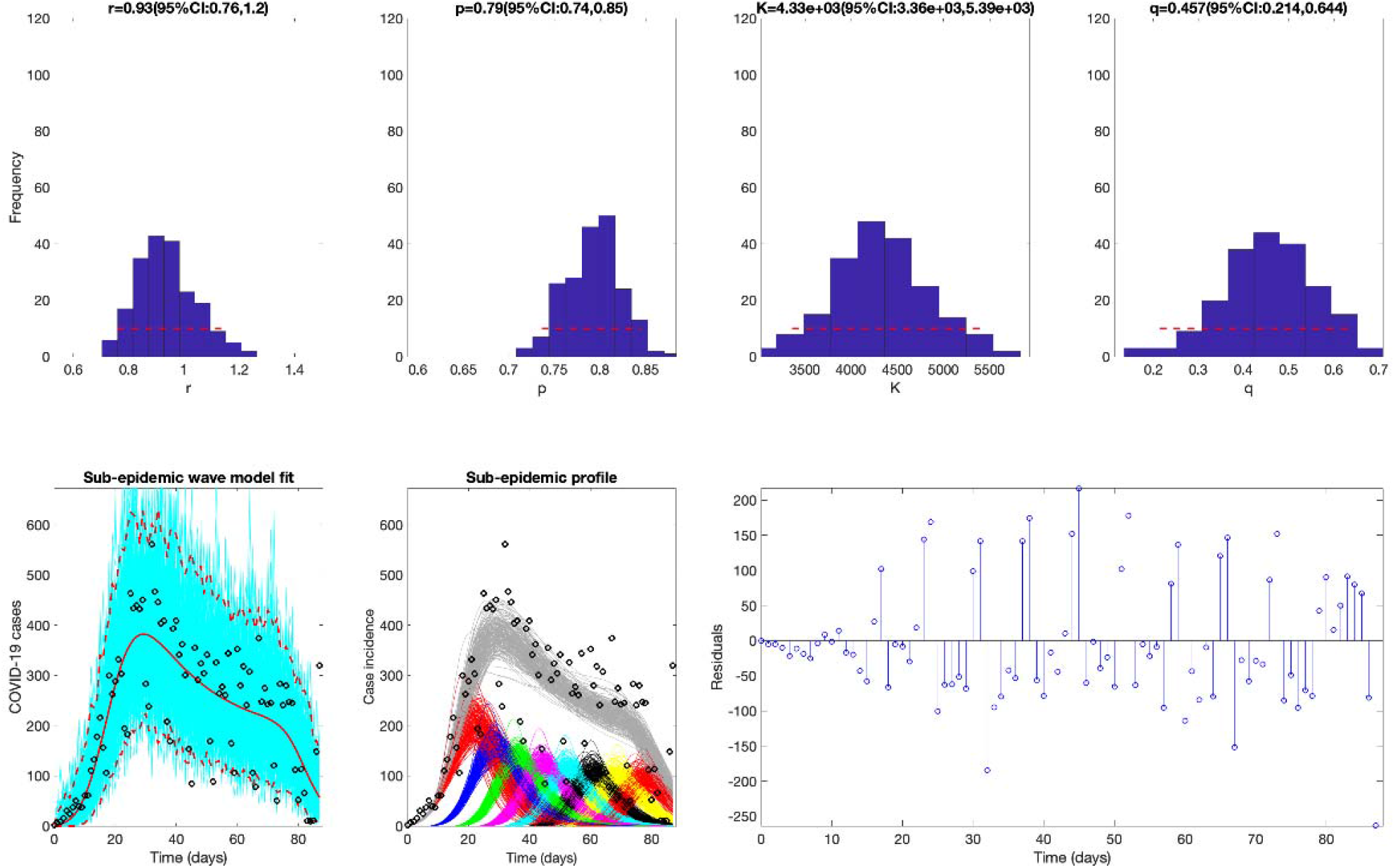
The best fit of the sub-epidemic model to the COVID-19 epidemic in Washington. The sub-epidemic wave model successfully captures the multimodal pattern of the COVID-19 epidemic. Further, parameter estimates are well identified, as indicated by their relatively narrow confidence intervals. The top panels display the empirical distribution of *r, p, K*, and *q*. Bottom panels show the model fit (left), the sub-epidemic profile (center), and the residuals (right). Black circles correspond to the data points. The best model fit (solid red line) and 95% prediction interval (dashed red lines) are also shown. Cyan curves are the associated uncertainty from individual bootstrapped curves. Three hundred realizations of the sub-epidemic waves are plotted using different colors.

**Figure S10.**
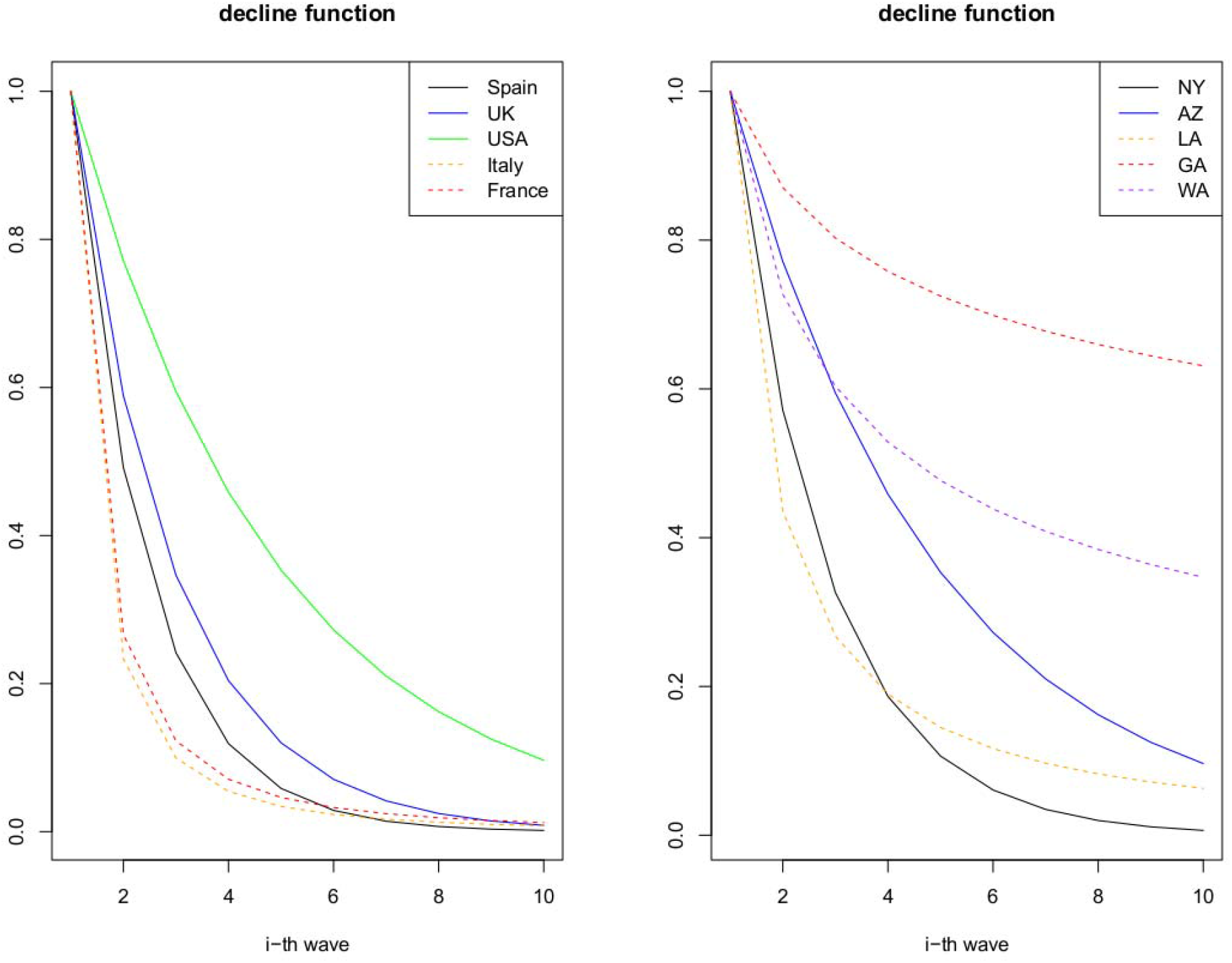
The sub-epidemic decline function across countries and USA states based on results presented in Table 1.

**Figure S11.**
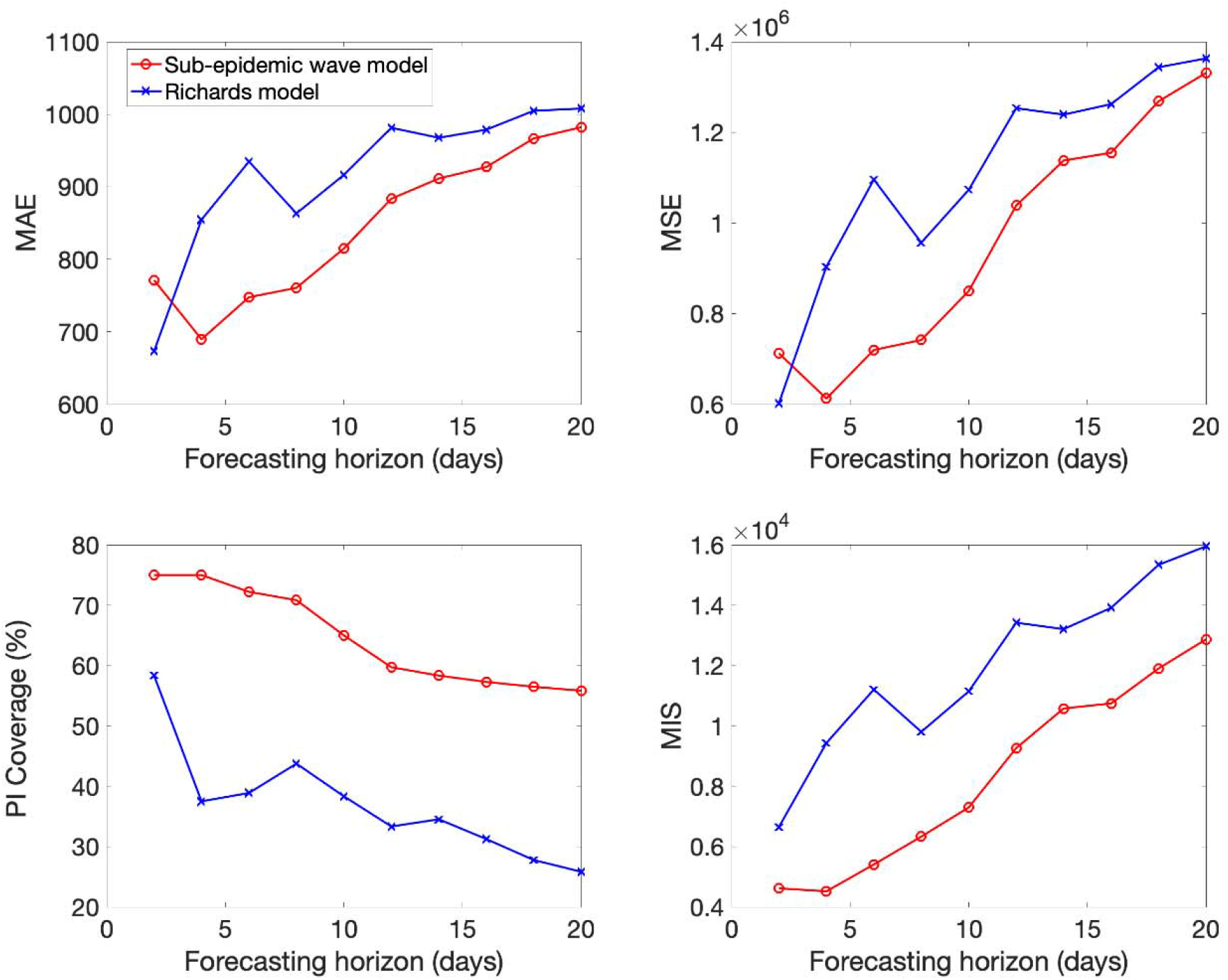
Mean performance of the sub-epidemic wave and the Richards models in 2-20 day ahead forecasts conducted during the epidemic in Italy. The sub-epidemic model outperformed the Richards model across all metrics and forecasting horizons except for 2-day ahead forecasts based on the MAE and the MSE.

**Figure S12.**
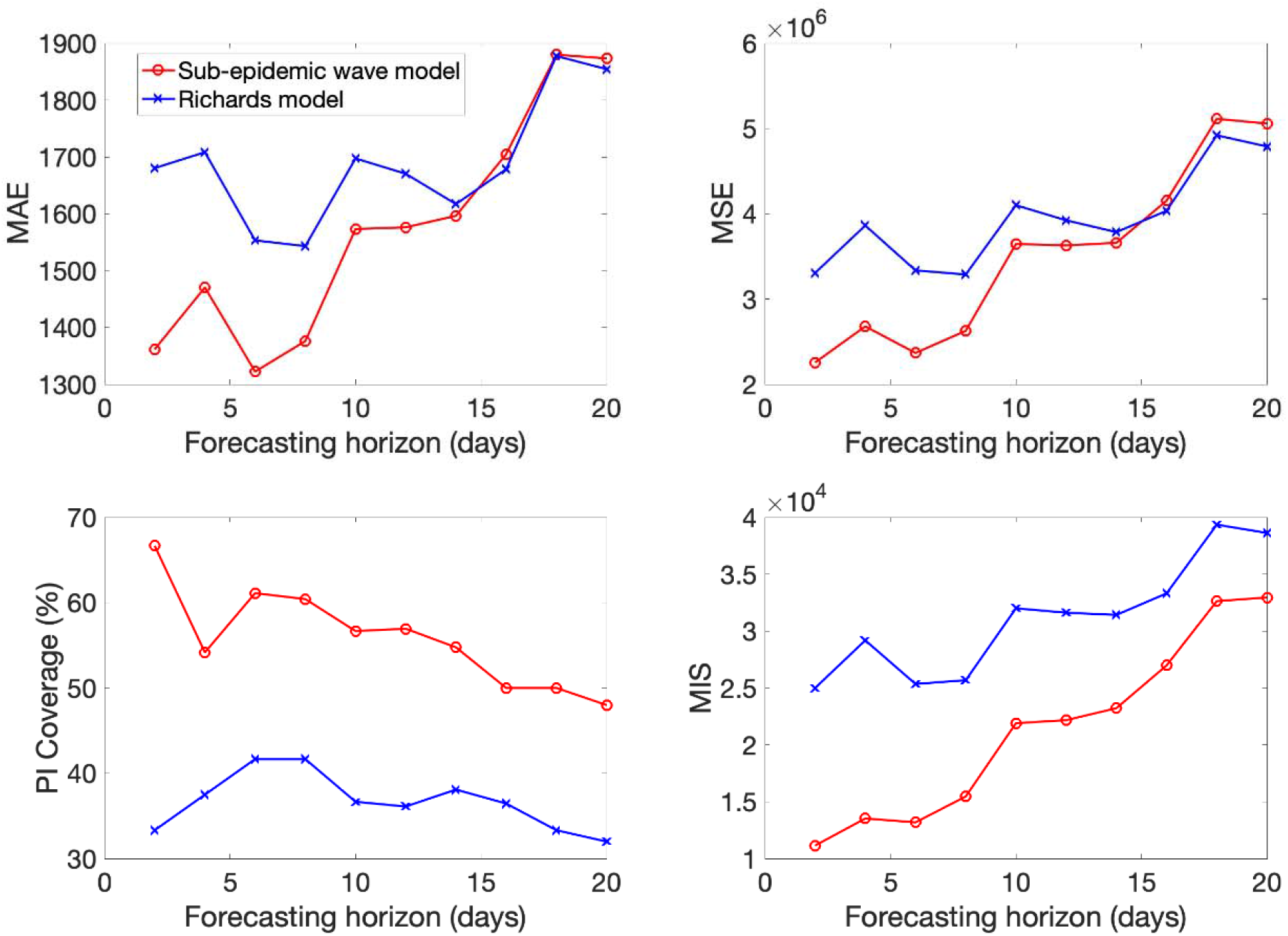
Mean performance of the sub-epidemic wave and the Richards models in 2-20 day ahead forecasts conducted during the epidemic in Spain. The sub-epidemic model outperformed the Richards model across all metrics and forecasting horizons, but the MSE and MAE reached similar values at longer forecasting horizons.

**Figure S13.**
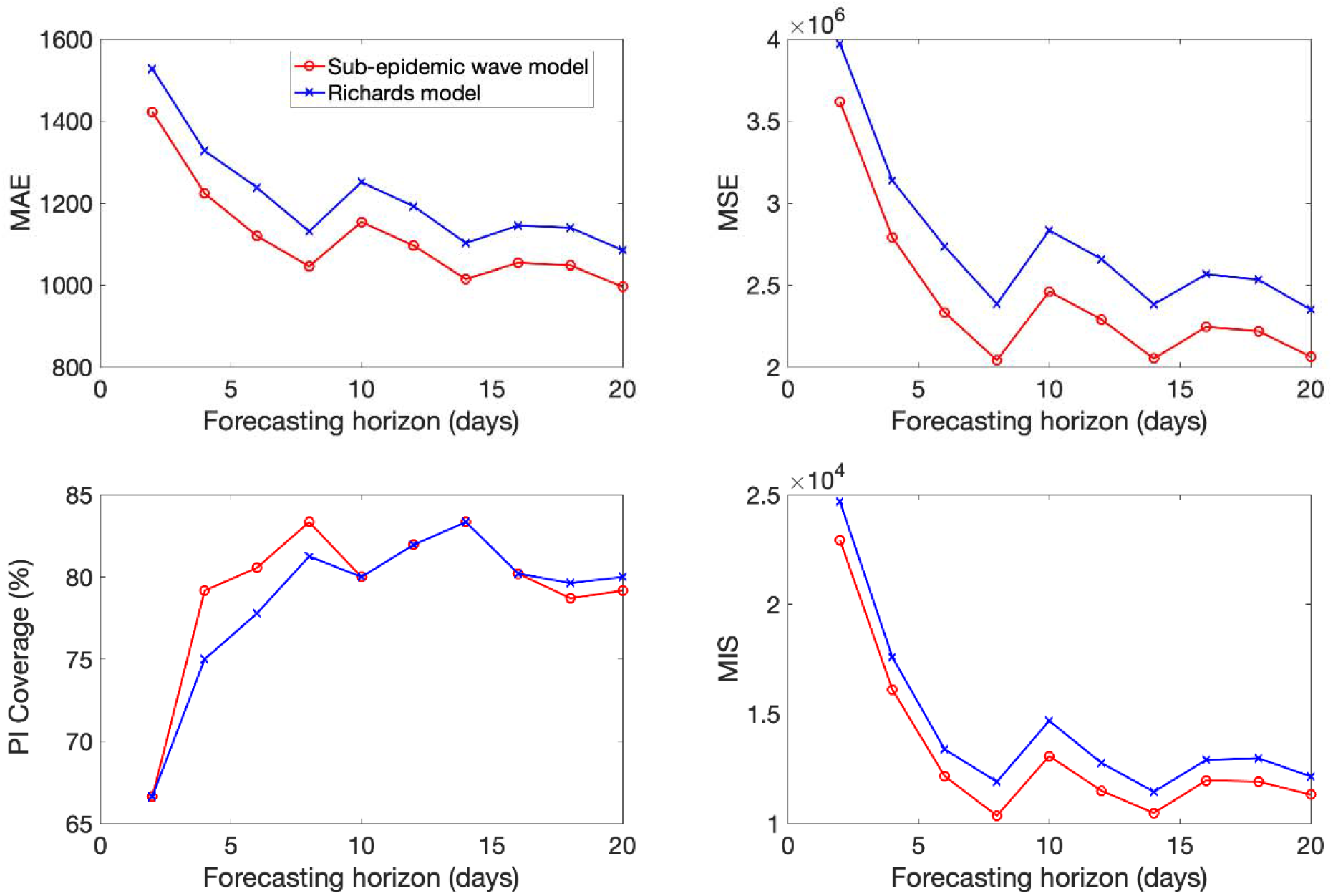
Mean performance of the sub-epidemic wave and the Richards models in 2-20 day ahead forecasts conducted during the epidemic in France. The sub-epidemic model outperformed the Richards model across all metrics and forecasting horizons.

**Figure S14.**
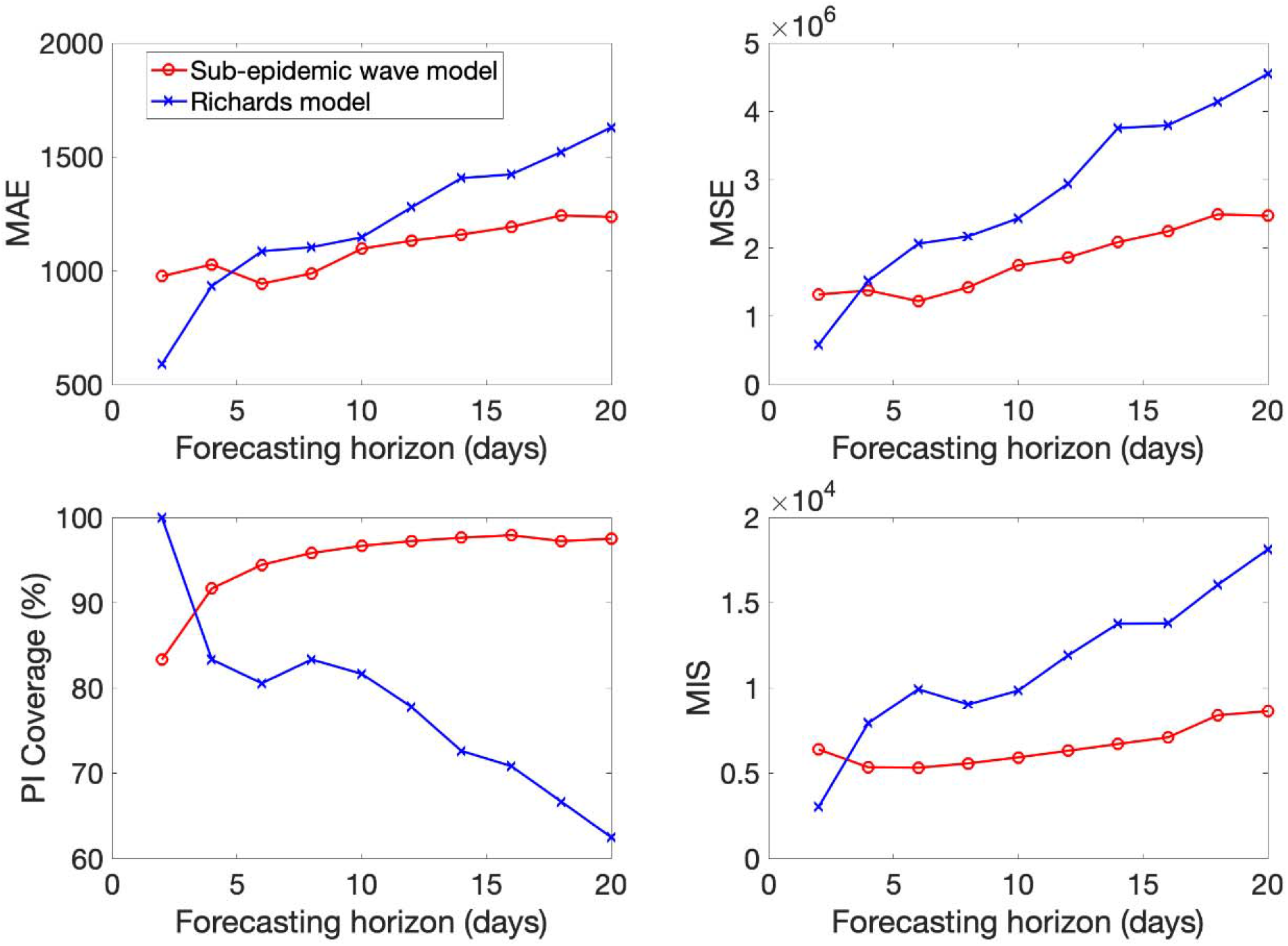
Mean performance of the sub-epidemic wave and the Richards models in 2-20 day ahead forecasts conducted during the epidemic in the UK. The sub-epidemic model outperformed the Richards model across all metrics and forecasting horizons except for 2-day ahead forecasts for which the Richards model reached somewhat better performance.

**Figure S15.**
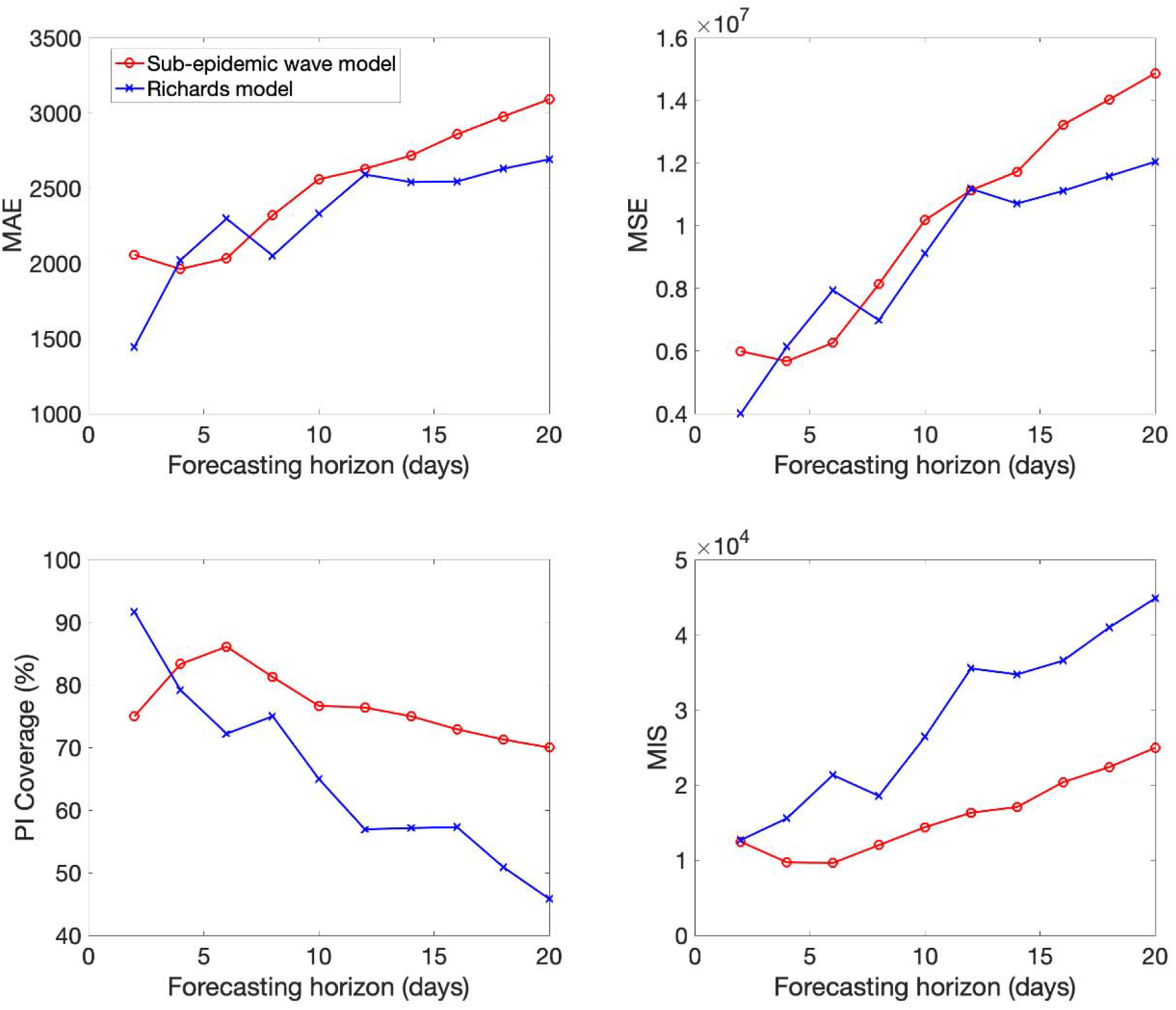
Mean performance of the sub-epidemic wave and the Richards models in 2-20 day ahead forecasts conducted during the epidemic in New York. The sub-epidemic model outperformed the Richards model across all forecasting horizons based on the PI Coverage and the MIS except for 2-day ahead forecasts. However, the Richards model more frequently outperformed the sub-epidemic wave model based on the MAE and MSE.

**Figure S16.**
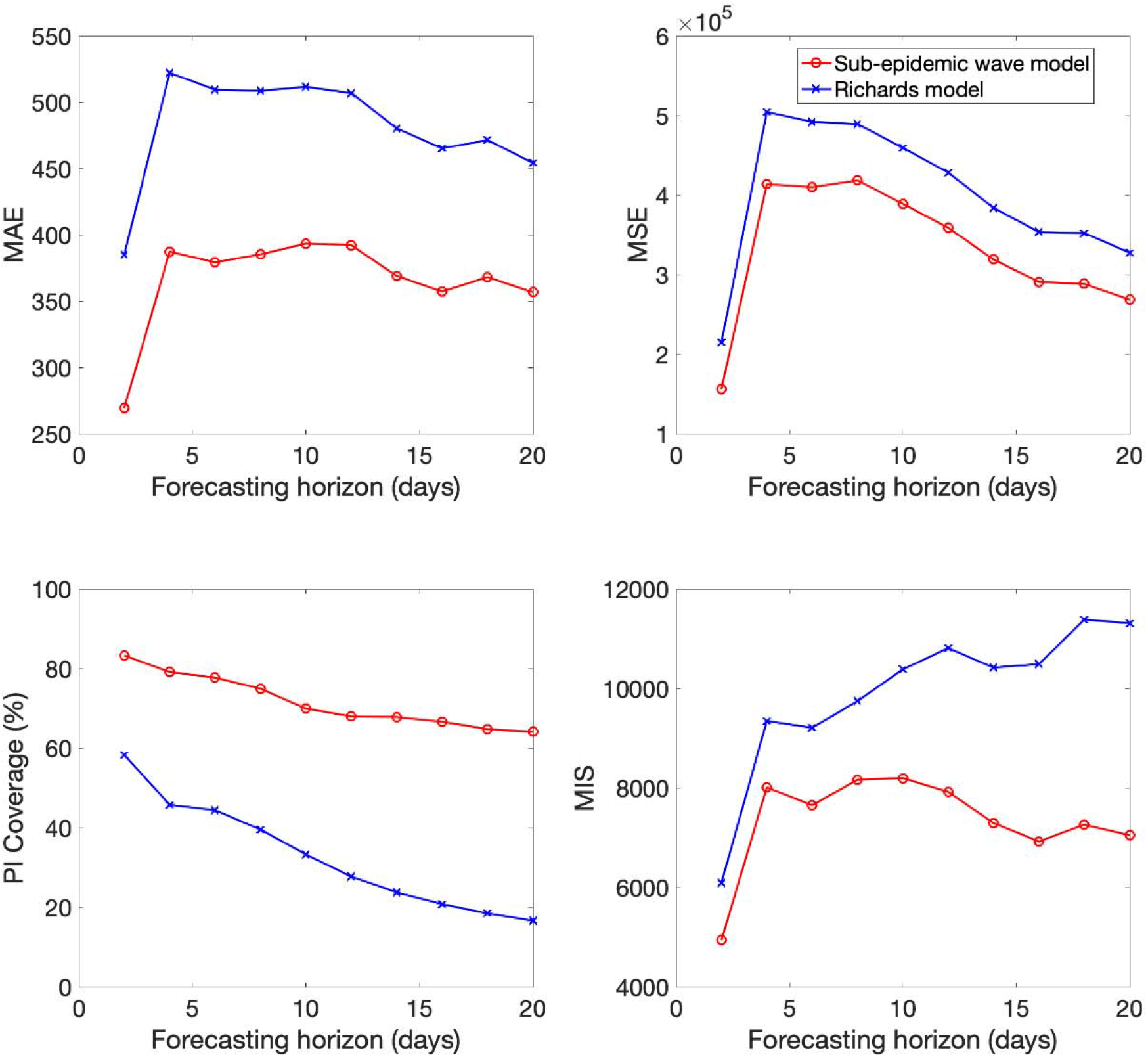
Mean performance of the sub-epidemic wave and the Richards models in 2-20 day ahead forecasts conducted during the epidemic in Louisiana. The sub-epidemic model outperformed the Richards model across all metrics and forecasting horizons.

**Figure S17.**
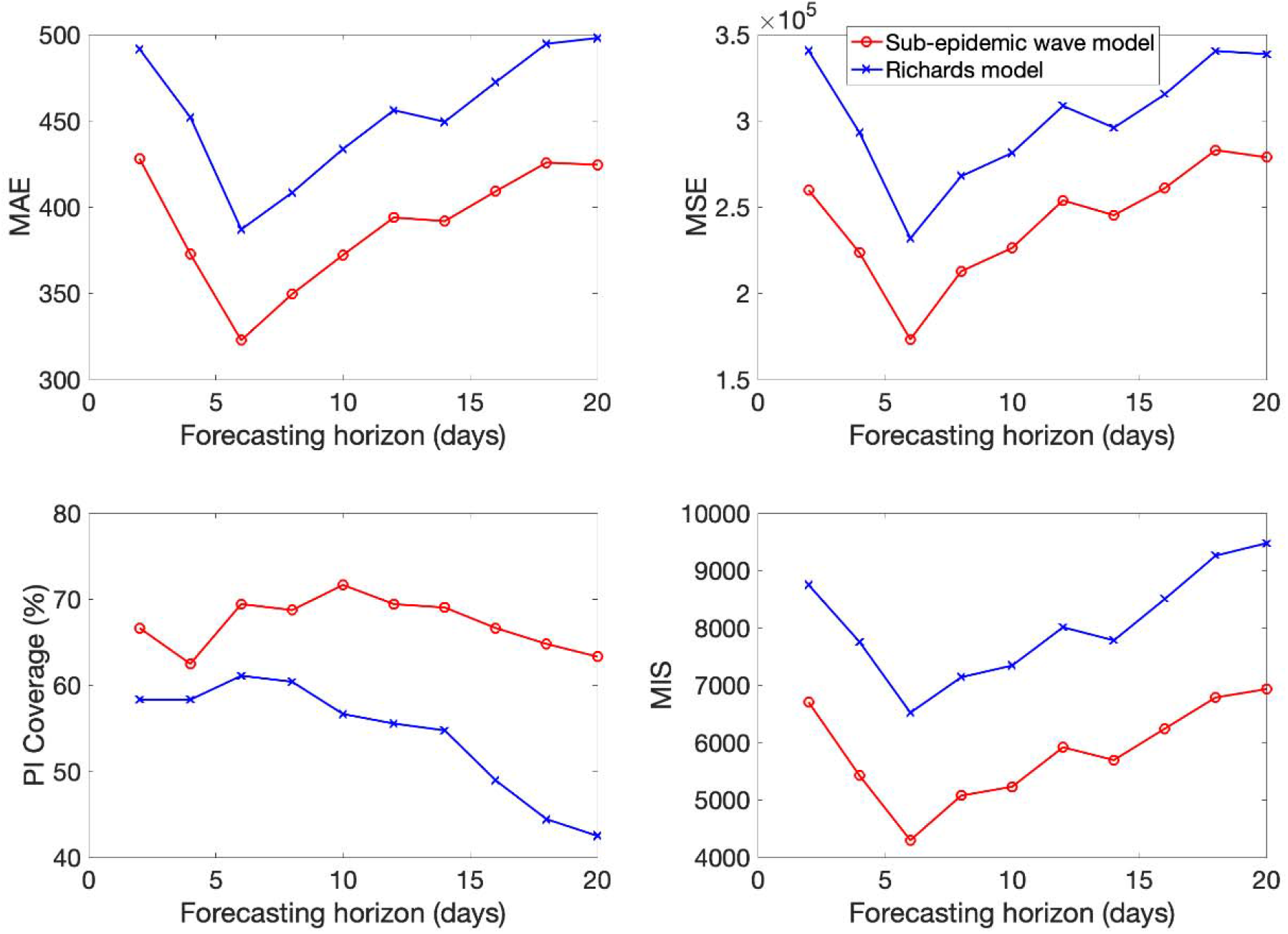
Mean performance of the sub-epidemic wave and the Richards models in 2-20 day ahead forecasts conducted during the epidemic in Georgia. The sub-epidemic model outperformed the Richards model across all metrics and forecasting horizons.

**Figure S18.**
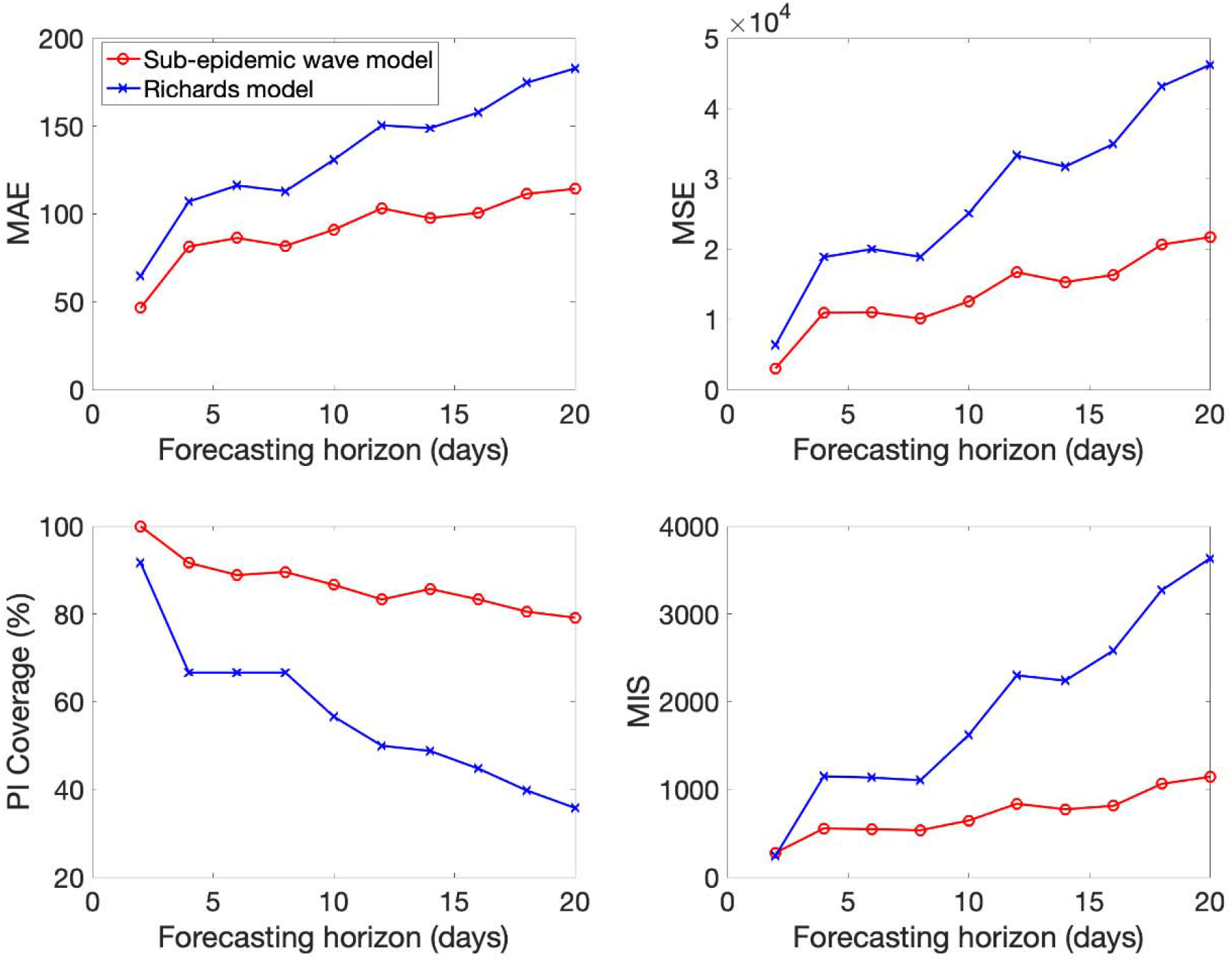
Mean performance of the sub-epidemic wave and the Richards models in 2-20 day ahead forecasts conducted during the epidemic in Arizona. The sub-epidemic model outperformed the Richards model across all metrics and forecasting horizons.

**Figure S19.**
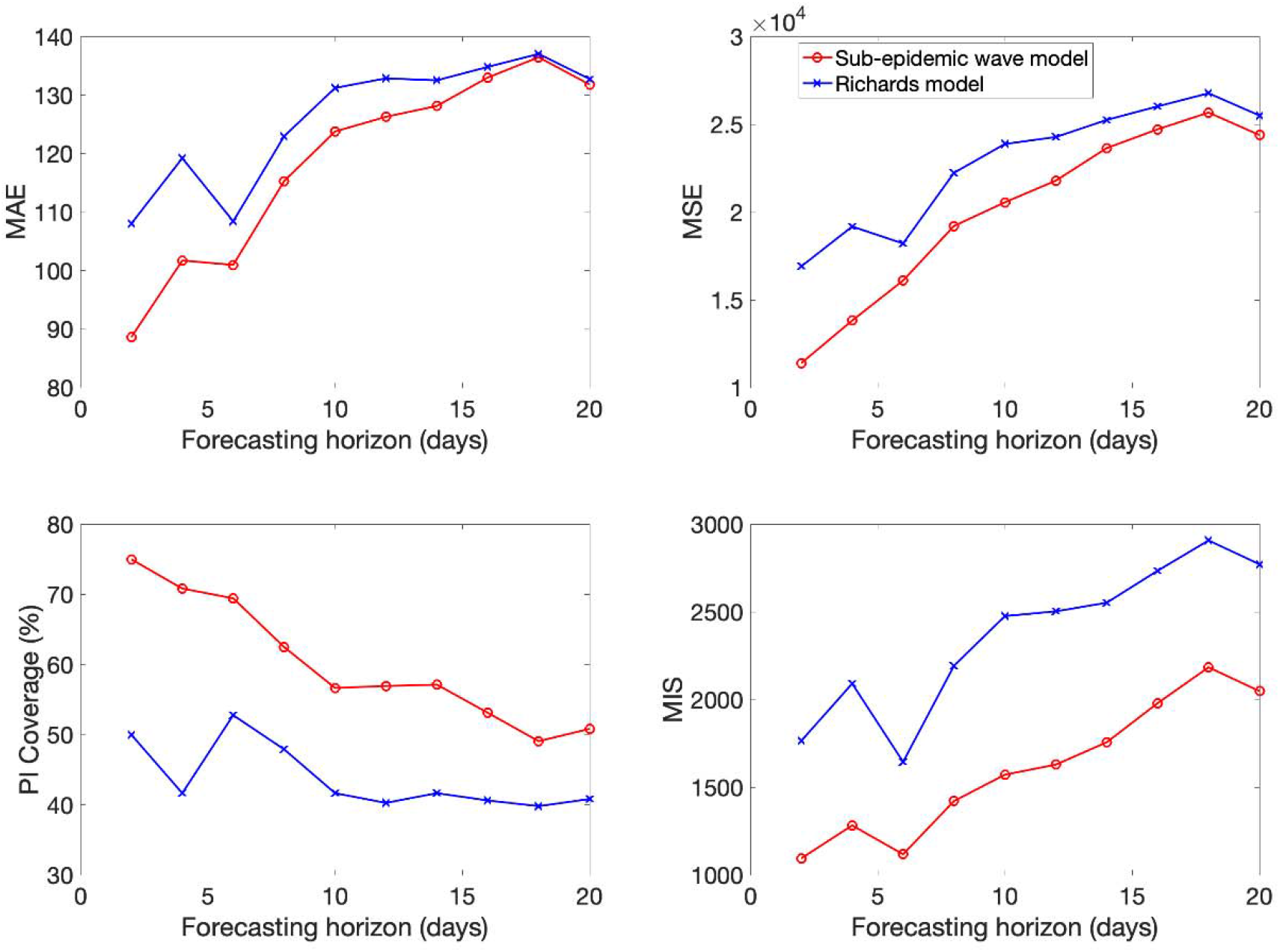
Mean performance of the sub-epidemic wave and the Richards models in 2-20 day ahead forecasts conducted during the epidemic in Washington. The sub-epidemic model outperformed the Richards model across all metrics and forecasting horizons.

**Figure S20.**
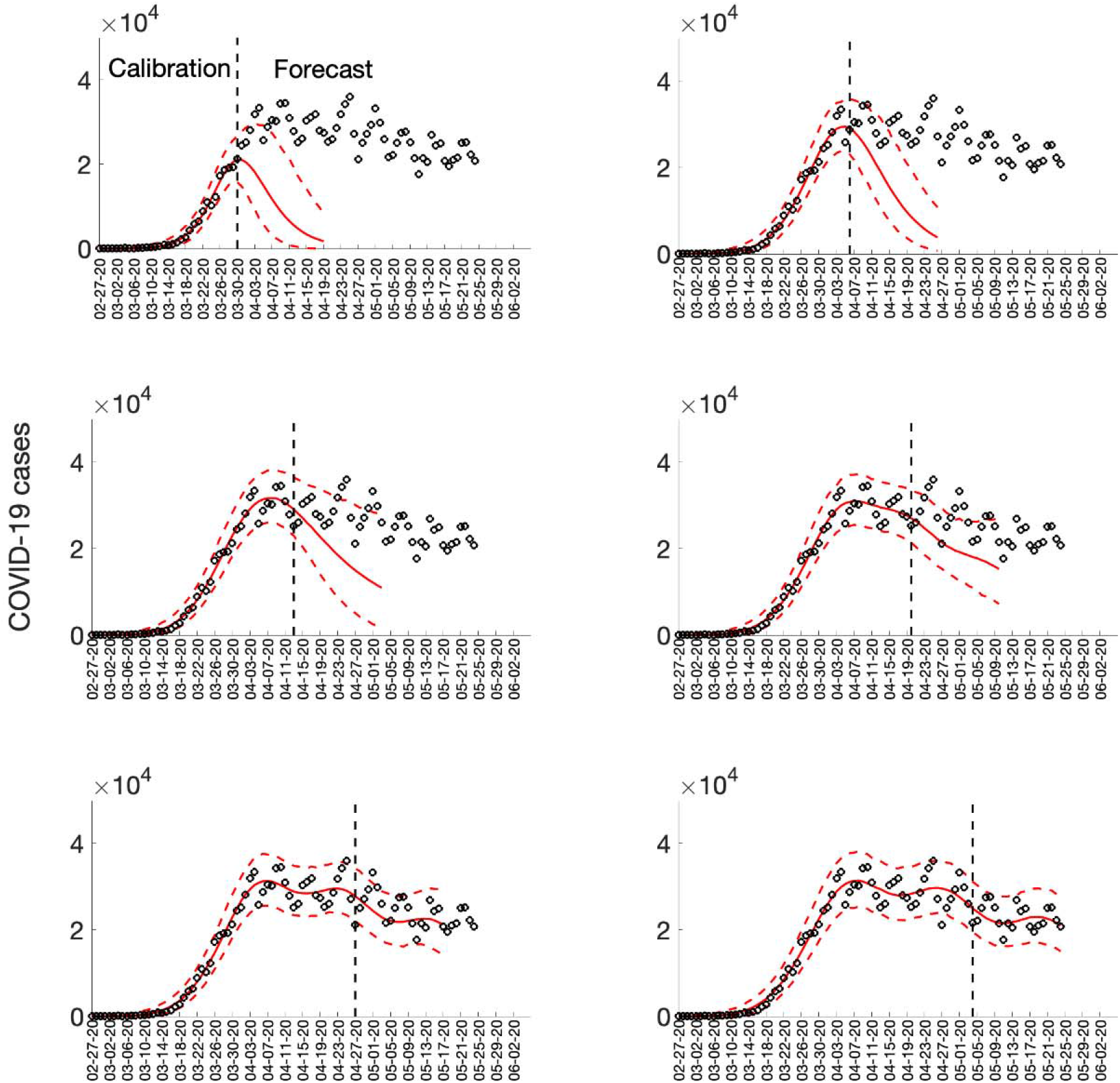
Sequential 20-day ahead forecasts of the sub-epidemic wave model for the COVID-19 epidemic in the USA. Black circles correspond to the data points. The model fit (solid red line) and 95% prediction interval (dashed red lines) are also shown. The vertical line separates the calibration period (left) from the forecasting period (right). The sequential forecasts were conducted on March 30, April 6, April 13, April 20, April 27, and May 4, 2020.

**Figure S21.**
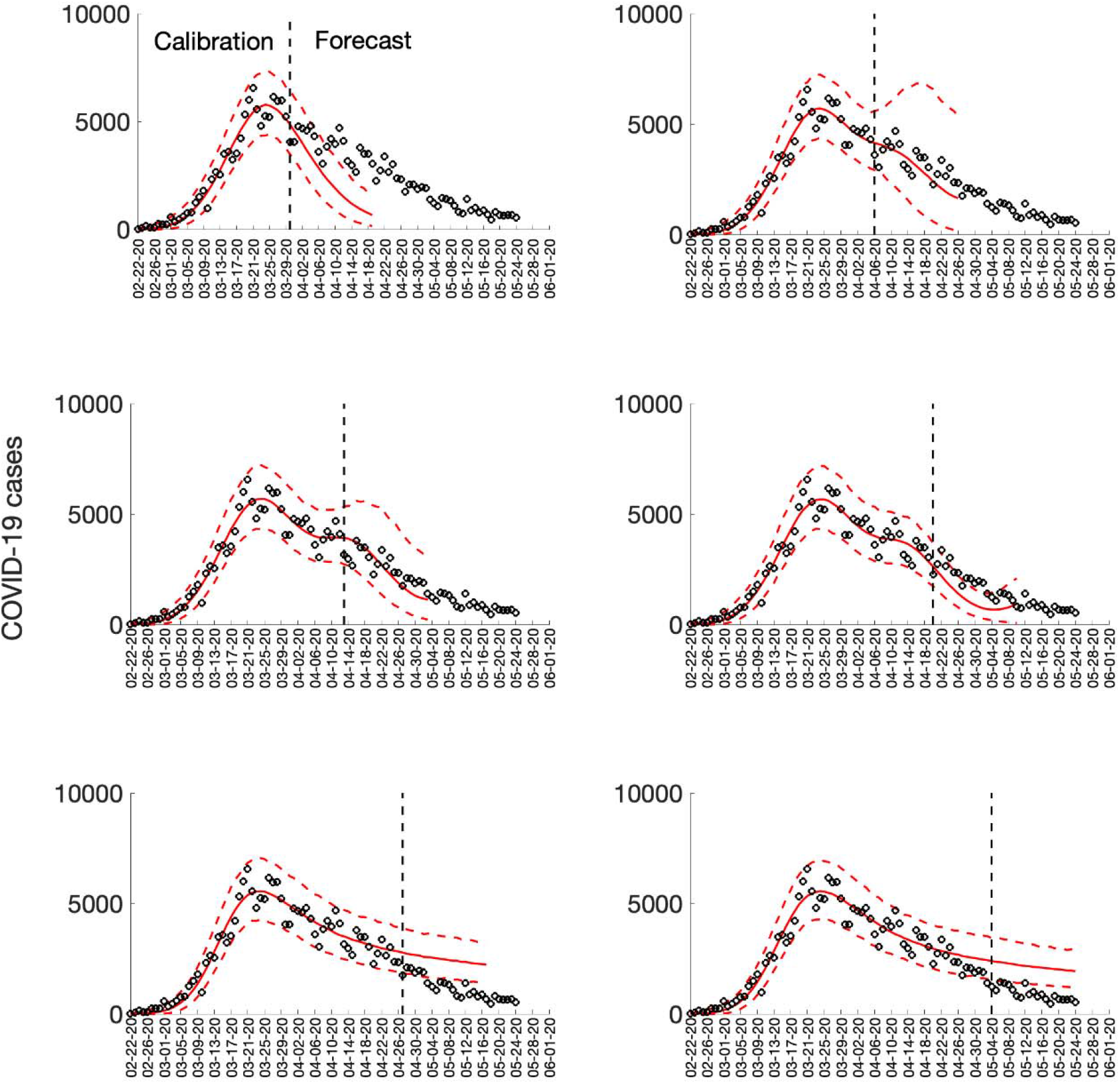
Sequential 20-day ahead forecasts of the sub-epidemic wave model for the COVID-19 epidemic in Italy. Black circles correspond to the data points. The model fit (solid red line) and 95% prediction interval (dashed red lines) are also shown. The vertical line separates the calibration period (left) from the forecasting period (right). The sequential forecasts were conducted on March 30, April 6, April 13, April 20, April 27, and May 4, 2020.

**Figure S22.**
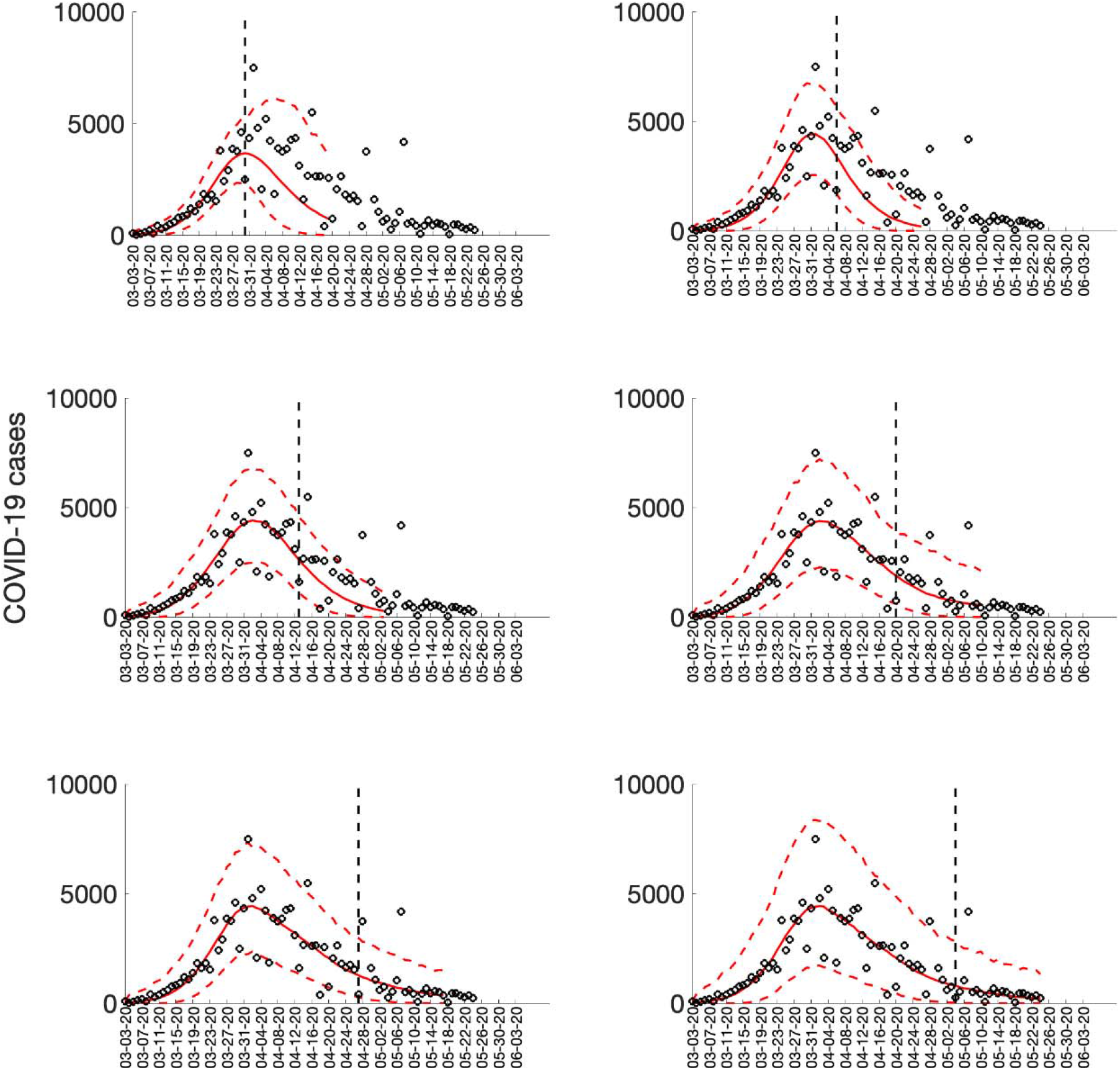
Sequential 20-day ahead forecasts of the sub-epidemic wave model for the COVID-19 epidemic in France. Black circles correspond to the data points. The model fit (solid red line) and 95% prediction interval (dashed red lines) are also shown. The vertical line separates the calibration period (left) from the forecasting period (right). The sequential forecasts were conducted on March 30, April 6, April 13, April 20, April 27, and May 4, 2020.

**Figure S23.**
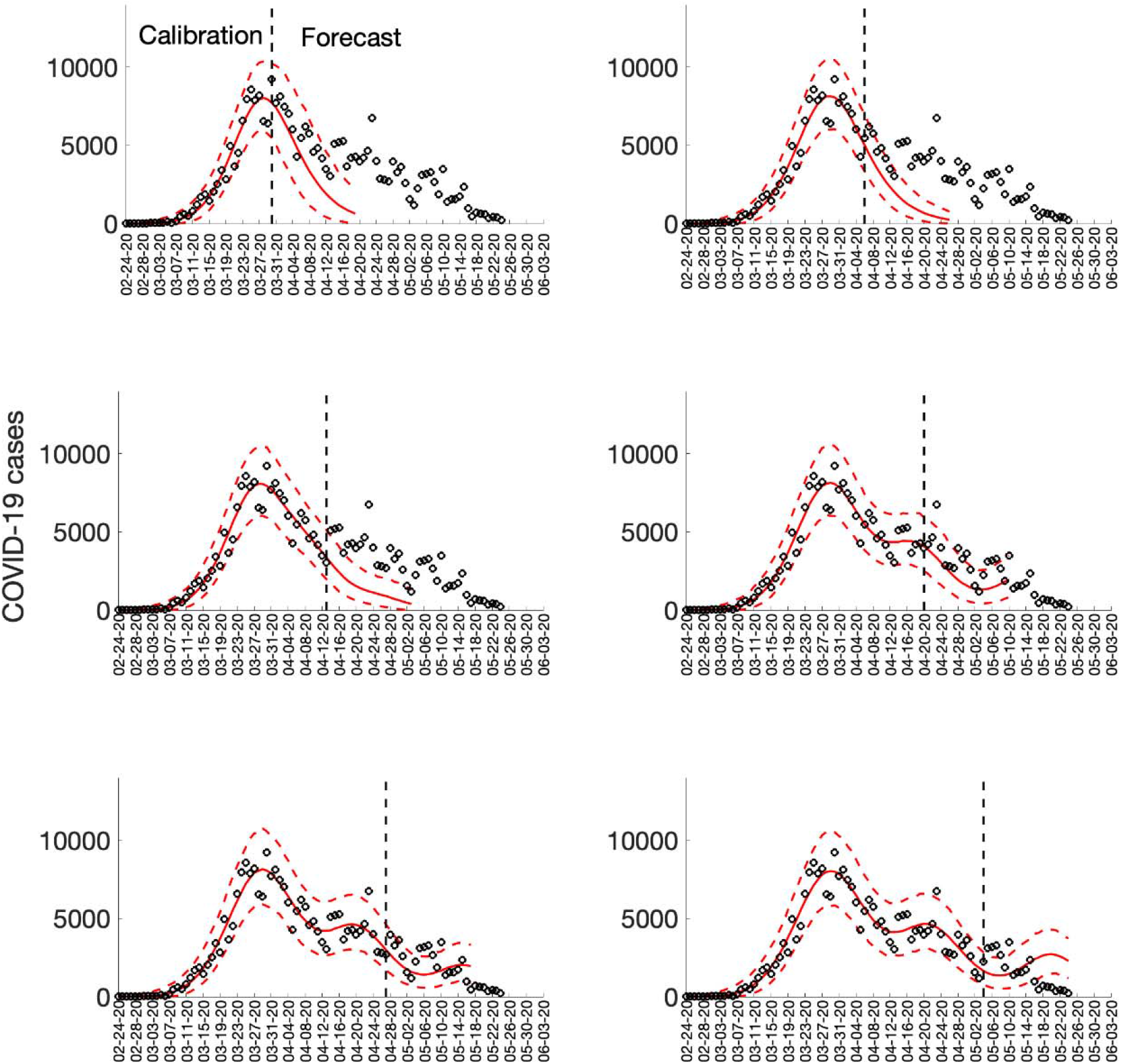
Sequential 20-day ahead forecasts of the sub-epidemic wave model for the COVID-19 epidemic in Spain. Black circles correspond to the data points. The model fit (solid red line) and 95% prediction interval (dashed red lines) are also shown. The vertical line separates the calibration period (left) from the forecasting period (right). The sequential forecasts were conducted on March 30, April 6, April 13, April 20, April 27, and May 4, 2020.

**Figure S24.**
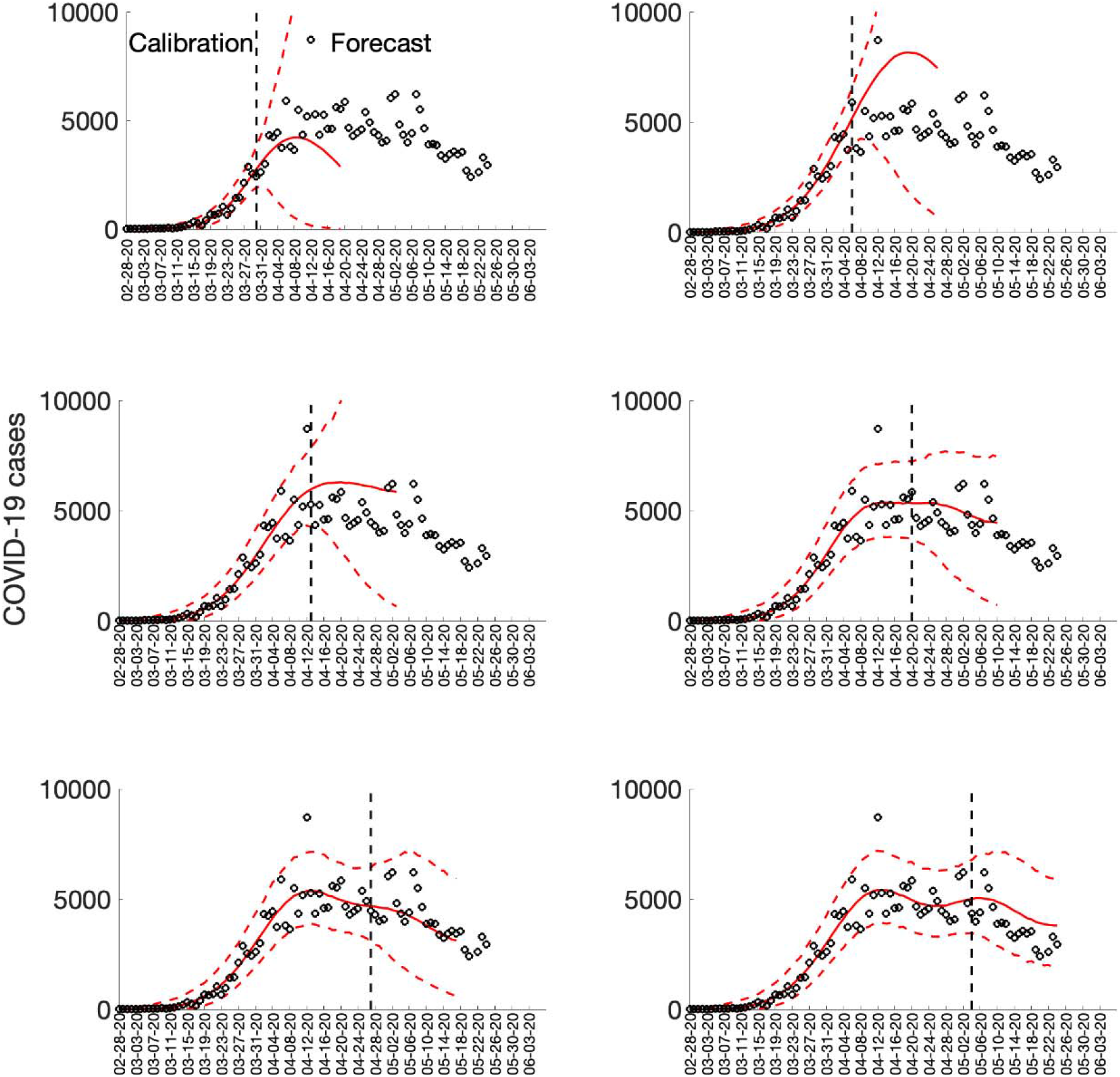
Sequential 20-day ahead forecasts of the sub-epidemic wave model for the COVID-19 epidemic in the UK. Black circles correspond to the data points. The model fit (solid red line) and 95% prediction interval (dashed red lines) are also shown. The vertical line separates the calibration period (left) from the forecasting period (right). The sequential forecasts were conducted on March 30, April 6, April 13, April 20, April 27, and May 4, 2020.

**Figure S25.**
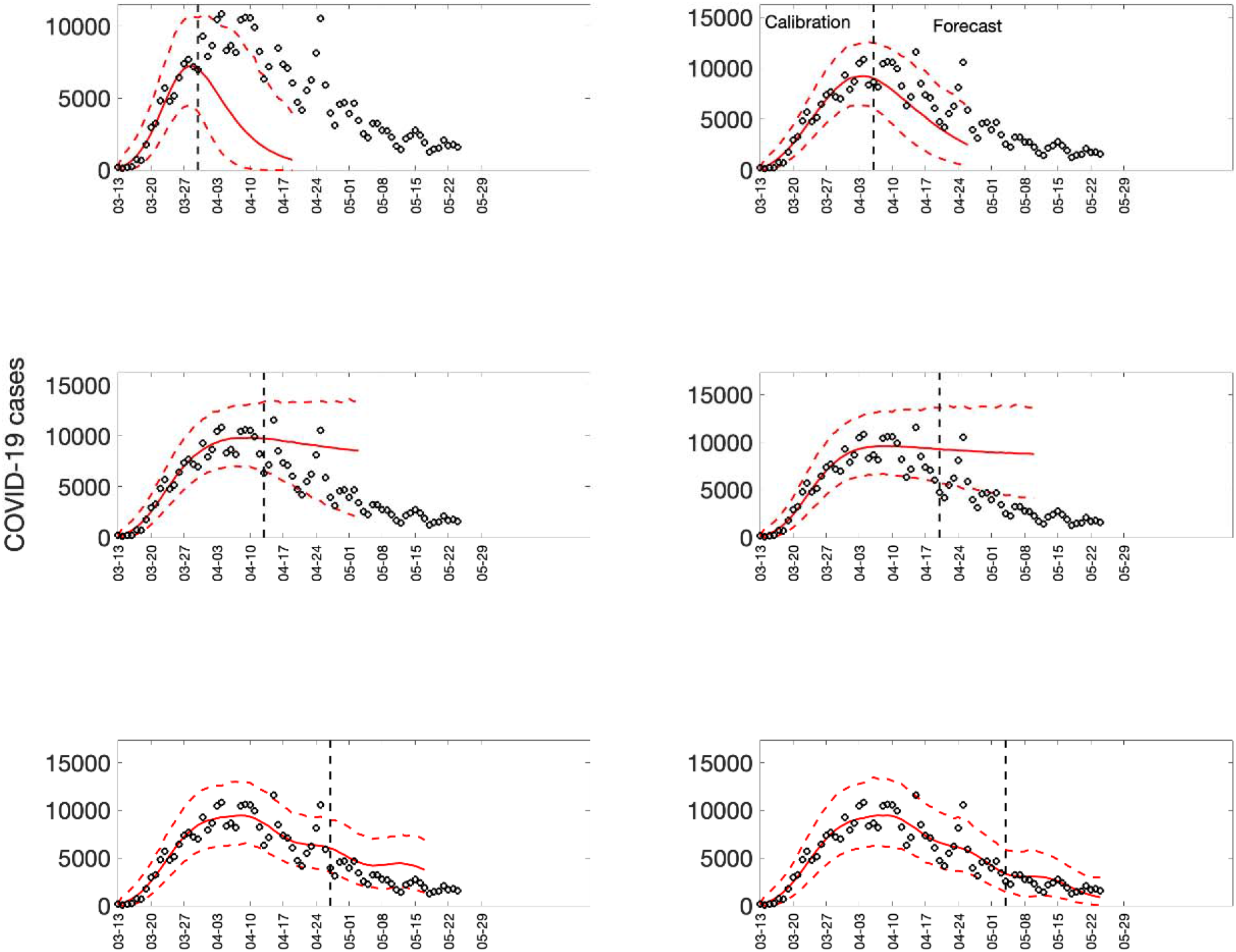
Sequential 20-day ahead forecasts of the sub-epidemic wave model for the COVID-19 epidemic in New York State. Black circles correspond to the data points. The model fit (solid red line) and 95% prediction interval (dashed red lines) are also shown. The vertical line separates the calibration period (left) from the forecasting period (right). The sequential forecasts were conducted on March 30, April 6, April 13, April 20, April 27, and May 4, 2020.

**Figure S26.**
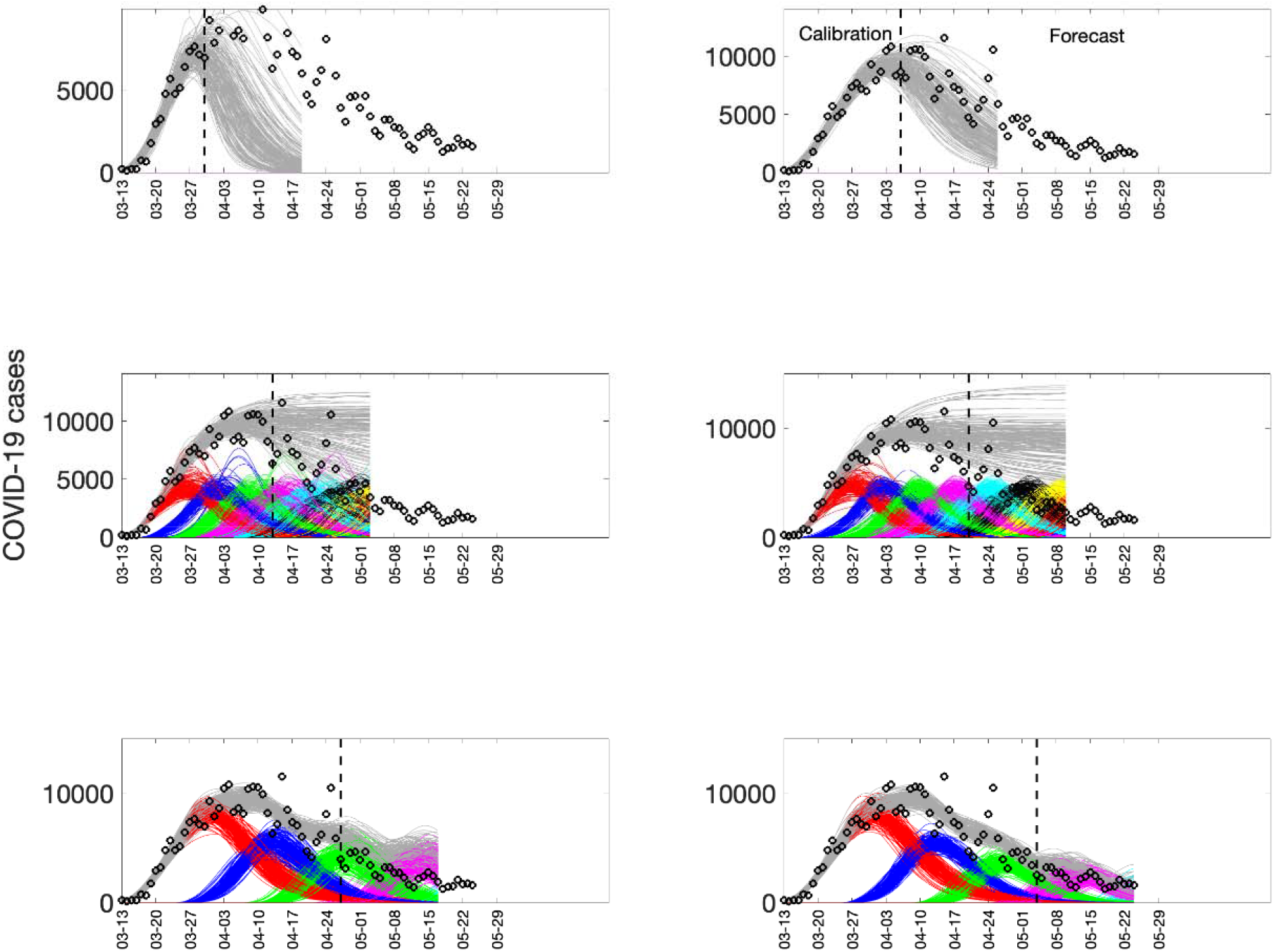
Sub-epidemic profiles of the sequential 20-day ahead forecasts for the COVID-19 epidemic in New York. Different colors represent different sub-epidemics of the epidemic wave profile. The aggregated trajectories are shown in gray and black circles correspond to the data points. The vertical line separates the calibration period (left) from the forecasting period (right). The sequential forecasts were conducted on March 30, April 6, April 13, April 20, April 27, and May 4, 2020.

**Figure S27.**
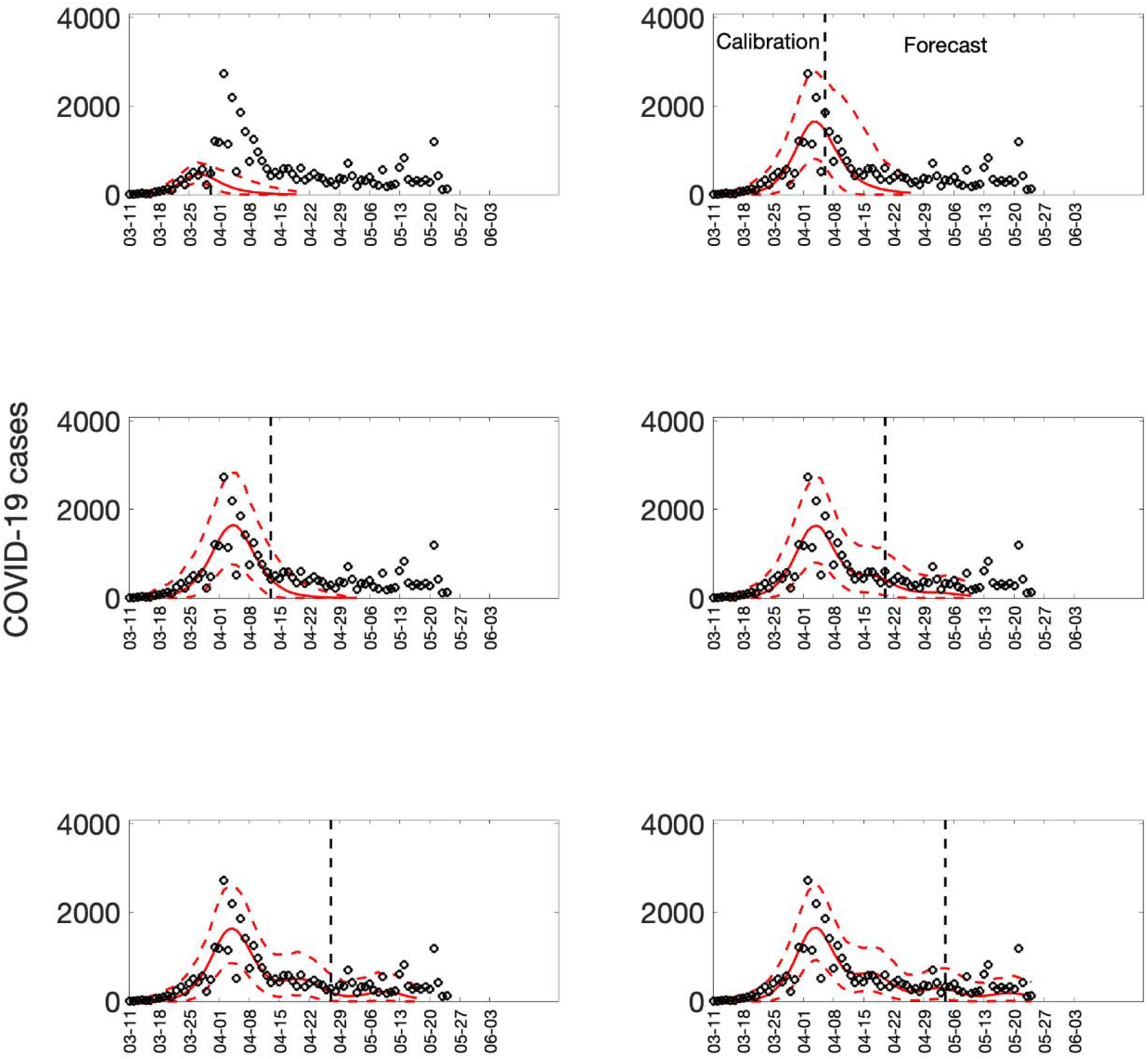
Sequential 20-day ahead forecasts of the sub-epidemic wave model for the COVID-19 epidemic in Louisiana. Black circles correspond to the data points. The model fit (solid red line) and 95% prediction interval (dashed red lines) are also shown. The vertical line separates the calibration period (left) from the forecasting period (right). The sequential forecasts were conducted on March 30, April 6, April 13, April 20, April 27, and May 4, 2020.

**Figure S28.**
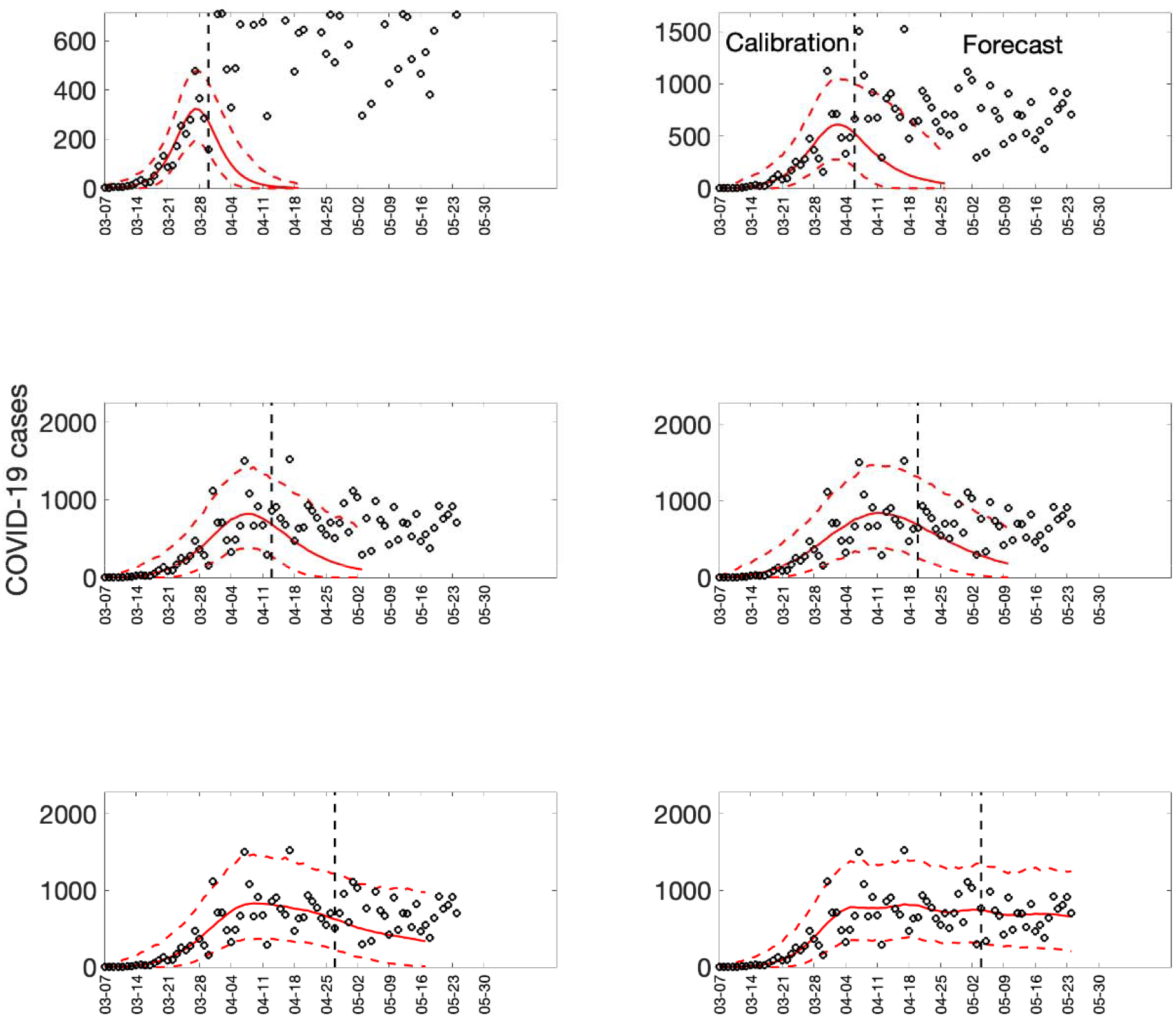
Sequential 20-day ahead forecasts of the sub-epidemic wave model for the COVID-19 epidemic in Georgia. Black circles correspond to the data points. The model fit (solid red line) and 95% prediction interval (dashed red lines) are also shown. The vertical line separates the calibration period (left) from the forecasting period (right). The sequential forecasts were conducted on March 30, April 6, April 13, April 20, April 27, and May 4, 2020.

**Figure S29.**
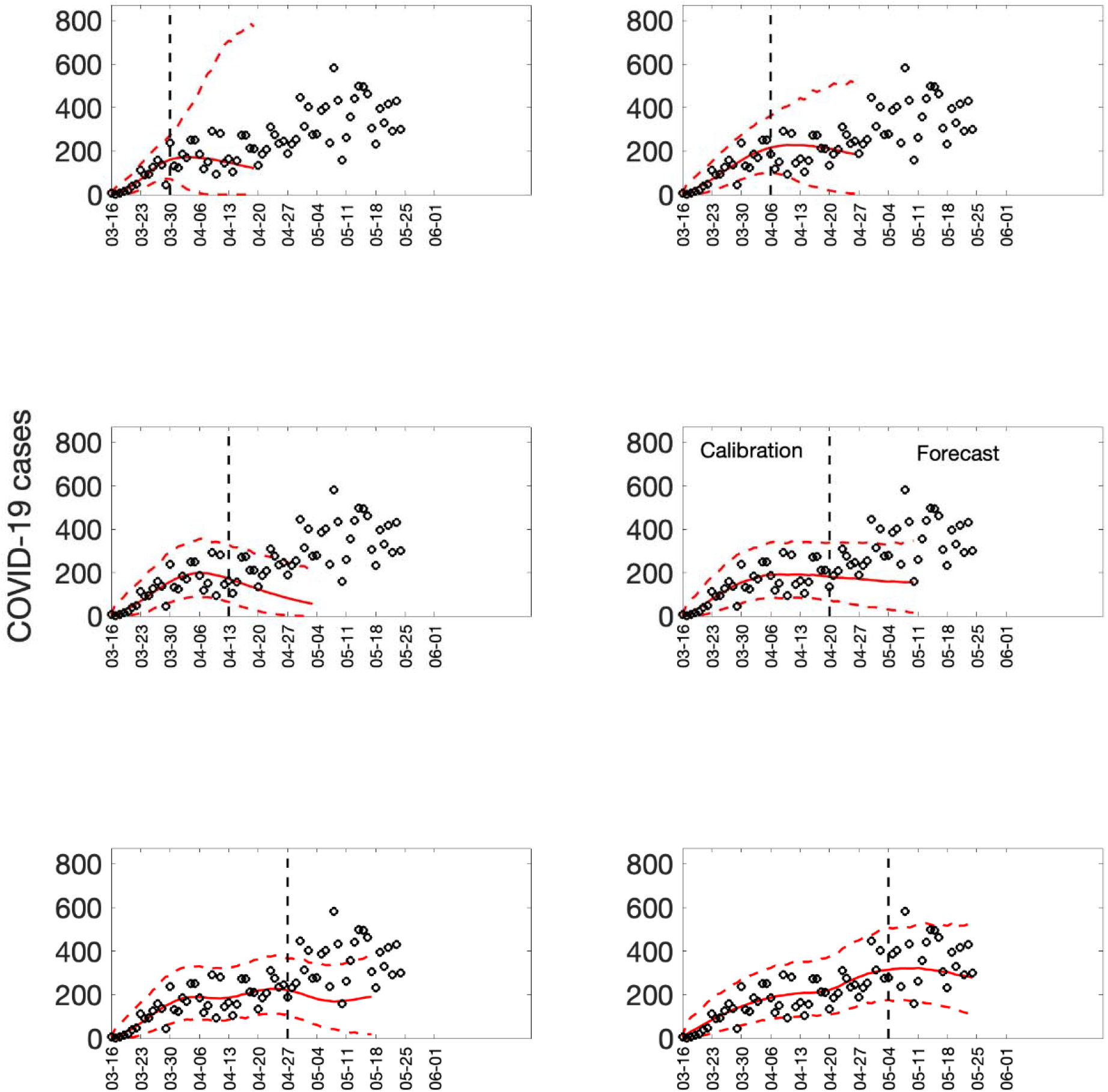
Sequential 20-day ahead forecasts of the sub-epidemic wave model for the COVID-19 epidemic in Arizona. Black circles correspond to the data points. The model fit (solid red line) and 95% prediction interval (dashed red lines) are also shown. The vertical line separates the calibration period (left) from the forecasting period (right). The sequential forecasts were conducted on March 30, April 6, April 13, April 20, April 27, and May 4, 2020.

**Figure S30.**
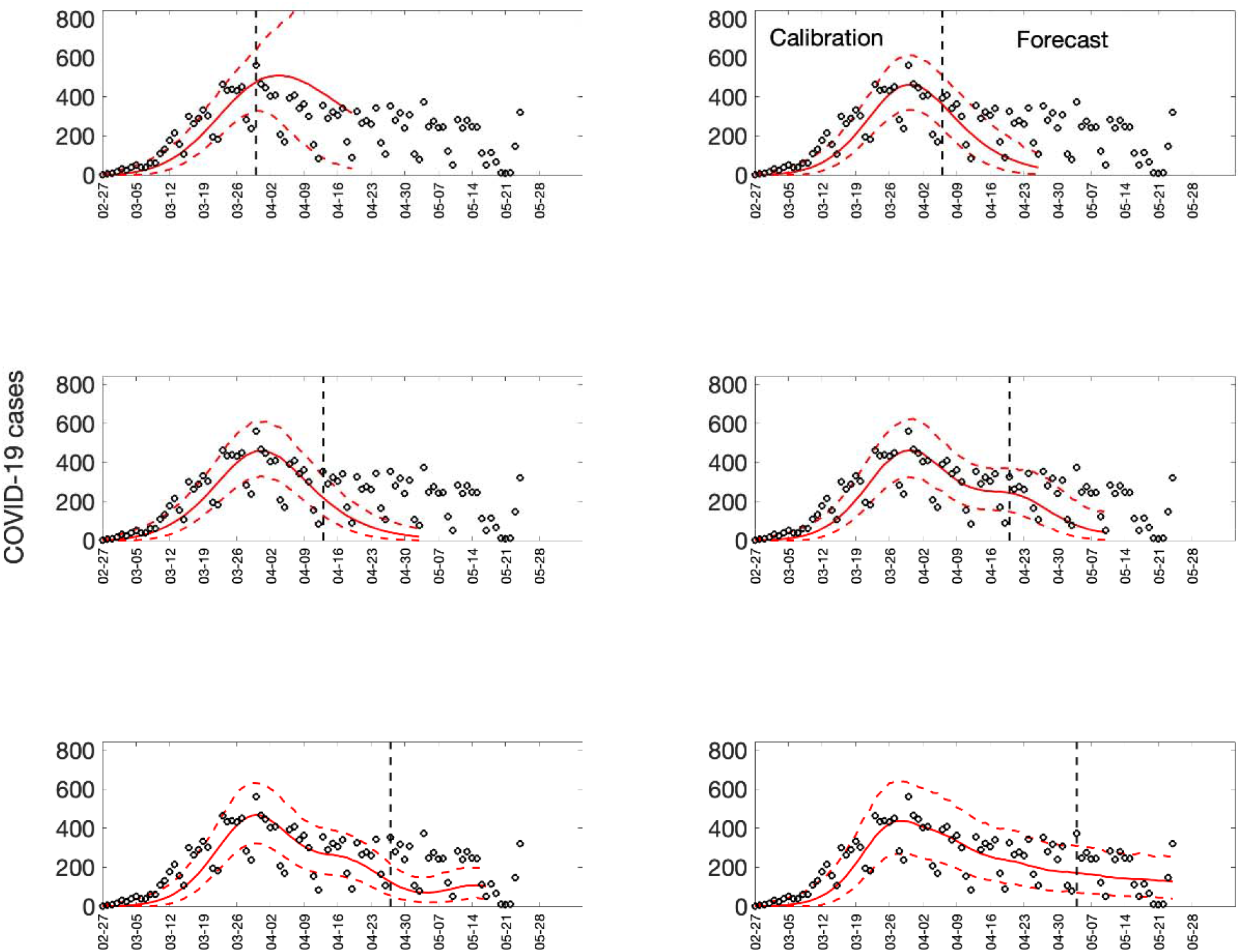
Sequential 20-day ahead forecasts of the sub-epidemic wave model for the COVID-19 epidemic in Washington. Black circles correspond to the data points. The model fit (solid red line) and 95% prediction interval (dashed red lines) are also shown. The vertical line separates the calibration period (left) from the forecasting period (right). The sequential forecasts were conducted on March 30, April 6, April 13, April 20, April 27, and May 4, 2020.

## References and Notes

1. Jewell NP, Lewnard JA, Jewell BL. Caution Warranted: Using the Institute for Health Metrics and Evaluation Model for Predicting the Course of the COVID-19 Pandemic. Annals of Internal Medicine 2020;173:xxx–xxx doi:107326/M20-1565.

2. Blower SM, McLean AR, Porco TC, et al. The intrinsic transmission dynamics of tuberculosis epidemics [see comments]. Nature Medicine 1995;95:815–21.

3. Garnett GP. The geographical and temporal evolution of sexually transmitted disease epidemics. Sexually Transmitted Infections 2002;78(Suppl 1):14–9.

4. Rothenberg R, Voigt R. Epidemiologic Aspects of Control of Penicillinase-Producing Neisseria gonorrhoeae. Sexually Transmitted Diseases 1988;15:211–6.

5. Rothenberg R, Dai D, Adams MA, Heath JW. The HIV endemic: maintaining disease transmission in at-risk urban areas. Sexually Transmitted Diseases 2017;44:71–8.

6. Chowell G, Tariq A, Hyman JM. A novel sub-epidemic modeling framework for short-term forecasting epidemic waves. BMC Med 2019;17:164-.

7. Wang XS, Wu J, Yang Y. Richards model revisited: validation by and application to infection dynamics. J Theor Biol 2012;313:12–9.

8. World Health Organization: Coronavirus disease (COVID-2019) situation reports. WHO. (Accessed May 15, 2020, at https://www.who.int/emergencies/diseases/novel-coronavirus-2019/situation-reports.)

9. Centro Nacional de Epidemiología (isciii): COVID-19 en España. (Accessed May 15, 2020, at https://cnecovid.isciii.es/.)

10. Github: Covid-19. (Accessed May 15, 2020, at https://github.com/pcm-dpc/COVID-19.) The COVID Tracking Project. (Accessed May 3, 2020, at https://covidtracking.com/data.)

12. Granovetter MS. The strength of weak ties. American Journal of Sociology 1973;78:1360–80.

12. Hsieh YH, Cheng YS. Real-time forecast of multiphase outbreak. Emerging infectious diseases 2006;12:122–7.

13. Worldometer. (Accessed May 11, 2020, at https://www.worldometers.info/coronavirus/.)

14. Banks HT, Hu S, Thompson WC. Modeling and inverse problems in the presence of uncertainty: CRC Press; 2014.

15. Chowell G, Ammon CE, Hengartner NW, Hyman JM. Transmission dynamics of the great influenza pandemic of 1918 in Geneva, Switzerland: Assessing the effects of hypothetical interventions. Journal of theoretical biology 2006;241:193–204.

16. Chowell G. Fitting dynamic models to epidemic outbreaks with quantified uncertainty: A Primer for parameter uncertainty, identifiability, and forecasts. Infect Dis Model 2017;2:379–98.

17. Chowell G, Tariq A, Hyman JM. A novel sub-epidemic modeling framework for short-term forecasting epidemic waves: Datasets and fitting code. figshare. Available from: http://10.6084/m9.figshare.8867882. 2019.

18. Gneiting T, Raftery AE. Strictly proper scoring rules, prediction, and estimation. J Am Stat Assoc 2007;102:359–78.

19. Kuhn M, Johnson K. Applied predictive modeling: New York: Springer; 2013.

20. M4Competition. Competitor’s Guide: Prizes and Rules. Available from: https://www.m4.unic.ac.cy/wp-content/uploads/2018/03/M4-Competitors-Guide.pdf (Accessed 04/01/2019).

21. Funk S, Camacho A, Kucharski AJ, Lowe R, Eggo RM, Edmunds WJ. Assessing the performance of real-time epidemic forecasts: A case study of Ebola in the Western Area region of Sierra Leone, 2014-15. PLoS computational biology 2019;15:e1006785.

## References

1. G. Chowell, A. Tariq, J. M. Hyman, A novel sub-epidemic modeling framework for short-term forecasting epidemic waves. BMC Med 17, 164–164 (2019).

2. H. T. Banks, S. Hu, W. C. Thompson, Modeling and inverse problems in the presence of uncertainty. (CRC Press, 2014).

3. G. Chowell, C. E. Ammon, N. W. Hengartner, J. M. Hyman, Transmission dynamics of the great influenza pandemic of 1918 in Geneva, Switzerland: Assessing the effects of hypothetical interventions. Journal of theoretical biology 241, 193–204 (2006).

4. G. Chowell, Fitting dynamic models to epidemic outbreaks with quantified uncertainty: A Primer for parameter uncertainty, identifiability, and forecasts. Infect Dis Model 2, 379–398 (2017).

5. G. Chowell, A. Tariq, J. M. Hyman, nd fitting code. figshare. Available from: http://10.6084/m9.figshare.8867882. (2019).

6. T. Gneiting, A. E. Raftery, Strictly proper scoring rules, prediction, and estimation. J Am Stat Assoc 102, 359–378 (2007).

7. M. Kuhn, K. Johnson, Applied predictive modeling. (New York: Springer, 2013), vol. 26.

8. M4Competition. Competitor’s Guide: Prizes and Rules. Available from: https://www.m4.unic.ac.cy/wp-content/uploads/2018/03/M4-Competitors-Guide.pdf (Accessed 04/01/2019).

9. S. Funk et al., Assessing the performance of real-time epidemic forecasts: A case study of Ebola in the Western Area region of Sierra Leone, 2014-15. PLoS computational biology 15, e1006785 (2019).

